# Systematic review of the effects of pandemic confinements on body weight and their determinants

**DOI:** 10.1101/2021.03.03.21252806

**Authors:** Moien AB Khan, Preetha Menon, Romona Govender, Amal Samra, Javaid Nauman, Linda Ostlundh, Halla Mustafa, Kholoud K Allaham, Jane E.M. Smith, Juma M Al Kaabi

## Abstract

Pandemics and subsequent lifestyle restrictions such as ‘lockdowns’ may have unintended consequences, including alterations in body weight. Understanding the impact and the mechanisms affecting body weight is paramount for planning effective public health measures for both now and future “lockdown”-type situations. This systematic review assesses and the impact of pandemic confinement on body weight and to identifies contributory factors. A comprehensive literature search was performed in seven electronic databases and in gray sources from their inception until 1^st^ July 2020 with an update in PubMed and Scopus on 1^st^ February 2021. In total, 2,361 unique records were retrieved, of which 41 studies were identified eligible: 1 case control study, 14 cohort and 26 cross-sectional studies (469, 362 total participants). The participants ranged in age from 6–86 years. The proportion of female participants ranged from 37% to 100%. Pandemic confinements were associated with weight gain in 7.2%–72.4% of participants and weight loss in 11.1%–32.0% of participants. Weight gain ranged from 0.6 (±1.3) to 3.0 (±2.4) kg, and weight loss ranged from 2.0 (±1.4) to 2.9 (±1.5) kg. Weight gain occurred predominantly in participants who were already overweight or obese. Associated factors included increased consumption of unhealthy food with decreased intake of healthy fresh fruits and vegetables, changes in physical activity, and altered sleep patterns. Weight loss during the pandemic was observed in individuals with previous low weight, and those who ate less and were more physically active before lock down. Associated factors included increased intake of fruits and vegetables, drinking more water and consuming no alcohol. Maintaining a stable weight was more difficult in populations with reduced income, particularly in individuals with lower educational attainment. The findings of this systematic review highlight the short-term effects of pandemic confinements. Learning from the “lockdown” experience is fundamental if we are to prepare for the next wave; a holistic, reactive, tailored response is needed involving multiple providers.

## Introduction

Devastating physical morbidity and mortality outcomes due to coronavirus disease 2019 (COVID-19) have been mitigated by ^(1,2)^ social distancing and quarantine measures ^(3)^, with significant direct and indirect health implications. Although lockdown has reduced the “R number”, physical wellbeing may have suffered from increased levels of stress, anxiety, and mental health issues ^(4–6)^. Moderate weight gain in people with a normal body mass index (BMI) has an adverse effect on metabolism, which increases the risk of diabetes, cardiovascular disease ^(7)^, or long-term ill-health ^(8)^. Lockdown may precipitate weight gain similar to that seen during the six-week summer holidays because of increased inactive time spent at home and snacking on energy dense foods ^(9–11)^. Rundle and colleagues argued that the extent and haste of the restrictions have exaggerated these observations ^(13)^ leading to rapid weight gain. This presents particular issues with the gained weight being more difficult to shed ^(14)^. Moreover, physical and social isolation is a recognized risk factor for obesity ^(15)^; with weight due to overconsumption, particularly when large “emergency” food stores are present ^(16)^. Reduced physical activity has further exacerbated the weight gain.

The COVID-19 outbreak adversely affected food supply and demand on a global scale ^(17)^. For some, lockdown gave more time to cook and overconsume, while those who were financially disadvantaged suffered from malnutrition and weight loss because of inflated food prices and food insecurity ^(18,19)^.

Recent research has linked obesity to an increased risk of contracting severe infections of COVID-19, thereby increasing the risk for extended hospitalization and increased mortality^(20)^. Importantly, therapeutic interventions and prophylactic treatments are more difficult and less effective in this group ^(21–26)^, with resultant poorer outcomes. Thus, weight gain secondary to pandemic confinement has an increased significance.

As the pandemic unfolds, researchers all over the globe try to better understand the prevalence, factors involved and impact of weight change in order to guide prevention strategies that will address this major public health crisis. These efforts have led to the identification of multiple determinants including biological, psychological and sociological processes that influence body weight during the pandemic. In this report, the interplay between these factors has been extrapolated from a systematic review of the current literature. Through an analysis of these observations, future public health interventions can be determined.

## Materials and Methods

### Methods and analysis

This review has been informed by the Cochrane Handbook for Systematic Reviews of Interventions ^(27)^ and is reported in accordance with the Preferred Reporting Guidelines for Systematic Reviews and Meta-analyses (PRISMA) ^(28)^. The review protocol is registered in the PROSPERO International Register of Systematic Reviews (Registration number CRD42020193440). This systematic review did not need approval from the ethics committee or required informed consent from the study populations as the data was retrieved from open-source databases and internet searches.

### Search strategy

A medical librarian (LÖ) performed a comprehensive literature search in the electronic databases PubMed Embase, Scopus, PsycInfo, Cochrane, CINAHL, and Web of Science in June and July of 2020. Search terms related to “pandemics” AND “body weight” AND “confinement” was systematically developed with the help of PubMed and PubMed’s MeSH and reviewed and discussed with a subject specialist (MK). The search string developed in PubMed was later adapted and applied to all databases. A combination of the search fields of “Title,” “Abstract,” and MeSH/Thesaurus (when available) was used to ensure the best possible search precision and results. No filters or limitations were applied to ensure the inclusion of pre-indexed materials. All databases were searched from their inception until July 2020. Selected sources of gray literature and the preprint archive medRxiv were additionally included in the literature search. A search update in PubMed and Scopus was conducted on February1, 2021. No additional relevant studies were located after hand screening the results from the updated search.

A search log with database specifications, detailed search strings, results, and notes for all sources included in the search is available in Appendix 1.

### Inclusion and exclusion criteria

All study designs relevant to human pandemic confinements and their effects on body weight were included (Table 1). All age groups were included, and there were no language restrictions.

**Table 1.** Inclusion and Exclusion Criteria.

|  | Inclusion | Exclusion |
| --- | --- | --- |
| **Population** | Human studies on pandemic confinement. | Animal studies.<br>Studies investigating the effect of obesity or overweight on various outcomes during the pandemics.<br>Diseases such as HIV, measles, and mumps. |
| **Effect** | Studies describing the impact of quarantine on body weight.<br>Studies showing the impact of quarantine on the overweight/obese population. | Studies showing obesity or overweight as a risk factor for the pandemic. |
| **Outcome** | Effect on body weight. Weight change (%), BMI change (kg/m<sup>2</sup>).<br>Demographic, behavioral, social, physical, psychological, lifestyle, and environmental behaviors during confinement that have effect bodyweight.<br>Studies showing measures taken to manage weight changes during confinement. |  |
| **Study** | Designs: All study designs. Language: All languages.<br>Year: Publication year inception – 1<sup>st</sup> February 2021. |  |

This review was extended to articles published from the time of inception until July 1^st^, 2020 and from an update in PubMed and Scopus on February 1^st^, 2021. The primary outcome was to determine the effects of pandemic confinements on body weight. The secondary research outcome was to identify factors affecting body weight during pandemic confinements.

We excluded animal studies and studies investigating the effect of obesity or overweight on various outcomes during the pandemic. We also excluded studies that only narrated the effects of obesity or overweight as a risk factor worsening pandemic-related disease. Studies on diseases, such as HIV, measles, and mumps, were also excluded.

### Screening and selection

All references identified in the databases and gray searches (n = 5,070) were uploaded to the systematic review tool Covidence (Veritas Health Innovation, 2020) for automatic deduplication and blinded screening (PRISMA flow diagram (Figure 1)). Two reviewers (HM and MK) independently screened the references at both the title/abstract (n= 2,361) and full-text level (n = 81). A third reviewer (PM) resolved any conflicts. The gray references and preprints were screened and deduplicated manually by MK and LÖ. Finally, the reference lists of the included papers were hand screened. Those full-text articles that did not meet the inclusion criterion were excluded (n = 27) (Figure 1). One study investigating weight gain exclusively in pregnant women was excluded ^(29)^ as it was impossible to distinguish physiological from pandemic-related weight gain in this group.

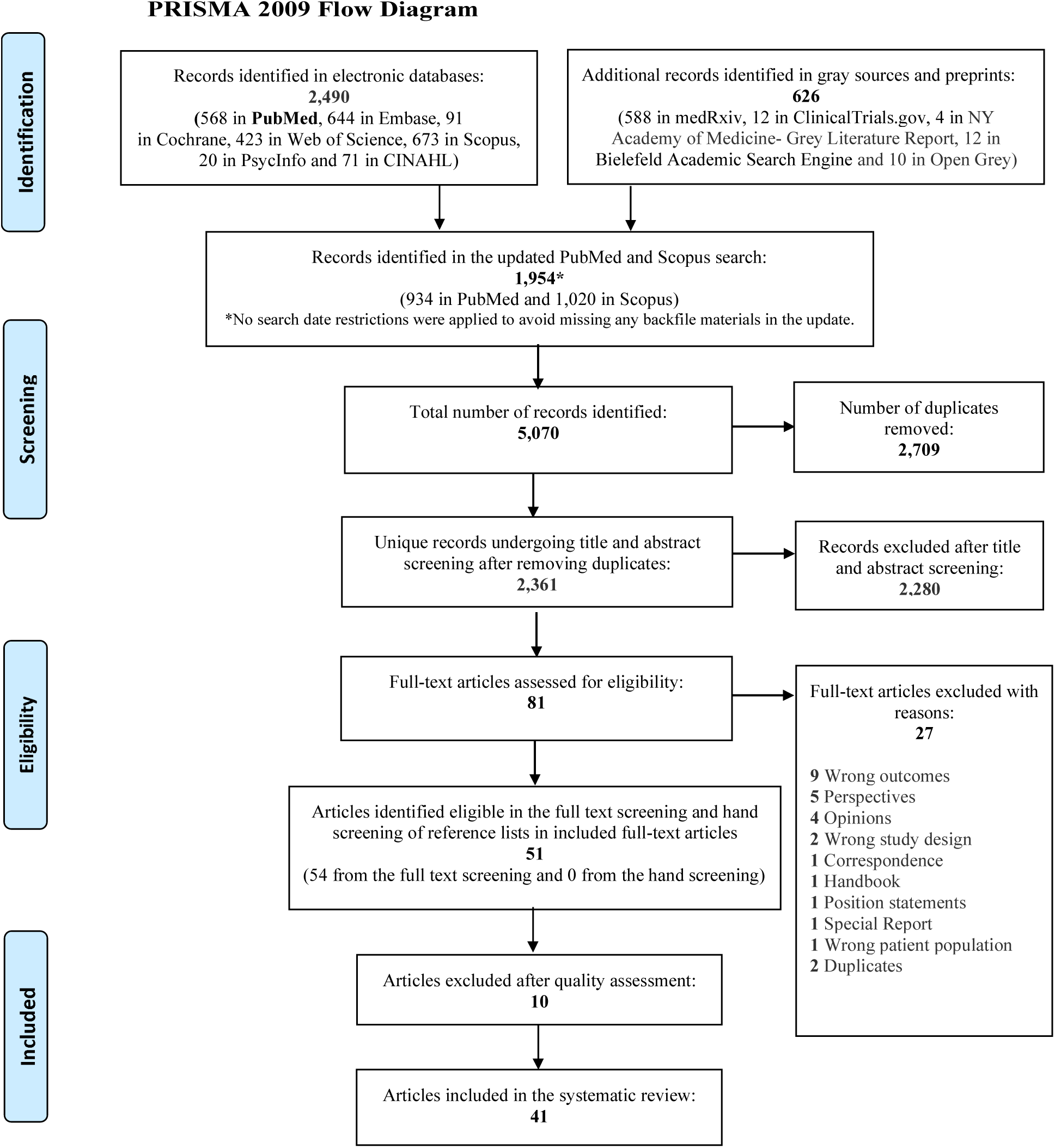

### Data extraction

The study characteristics including the authors, year of publication, country of origin, study design, research instruments used, validity of survey questionnaire, proportion of female participants, age range and mean age of participants, mean BMI of participants, and mean weight of participants were extracted by one reviewer (MK). The other reviewers (PM, RG, and AS) extracted and reviewed the data independently. Determinants that had an impact on body weight were extracted and reviewed (primarily by MK and secondarily by PM, RG, and AS).

### Quality assessment (n = 51)

Two reviewers (MK and PM) independently performed a quality assessment of the 54 studies identified as eligible in the screening (Appendix 2). We applied a validated Newcastle–Ottawa Quality Assessment Scale to assess the quality of the studies that were included in the review ^(30–32)^. Quality scores obtained via the Newcastle–Ottawa scale for cross-sectional, cohort studies and case-control studies were used to assess selection, comparison, and outcomes. Score disagreements were resolved through a discussion between MK and PM, and a final consensual rating was assigned to each study. Studies six or more stars were considered high quality and were included in the review. Studies with fewer than six stars were excluded (Appendix 2).

## Results (n = 41)

### Categorization of determinants

Ten studies met the inclusion criteria covering pandemic confinements and their effects. These were then further subdivided into the following five main categories:

a. Demographic determinants
b. The impact of pandemic confinements on body weight
c. Dietary changes and other lifestyle behavior changes during the confinement
d. Behavior changes observed in obese participants
e. Determinants of obesity during pandemic confinements

Our search yielded 5,070 records of which 2,361 unique studies remained after de-duplication. After applying the inclusion and exclusion criteria in the title and abstract screening, 81 articles were eligible for full-text screening (Figure 1). We excluded 10 studies based on a quality assessment of the results, and 27 studies were excluded based on reasons presented in the PRISMA flow diagram (Figure 1). The range of observations covered dietary choices ^(14,21–26,33–36,36–47)^, lifestyle changes in children ^(24,36,48–51)^, physical activity levels ^(33–35,37,38,38–41,43,46,47,49,52,53,53–60)^, psychosocial factors^(22,23,26,38,44,45,51,52,55,56,58,61,62)^, socioeconomic factors ^(23,37,48,52,54,61)^ and sleeping patterns ^(26,46,51,63)^.

### Demographic determinants (study and sample characteristics) (n = 41)

Table 2 describes the characteristics of each of the 41 included studies. All of the studies were published in 2020 and 2021. Two studies were from preprints and were included after assessing their qualities individually ^(22,52)^.

**Table 2:**
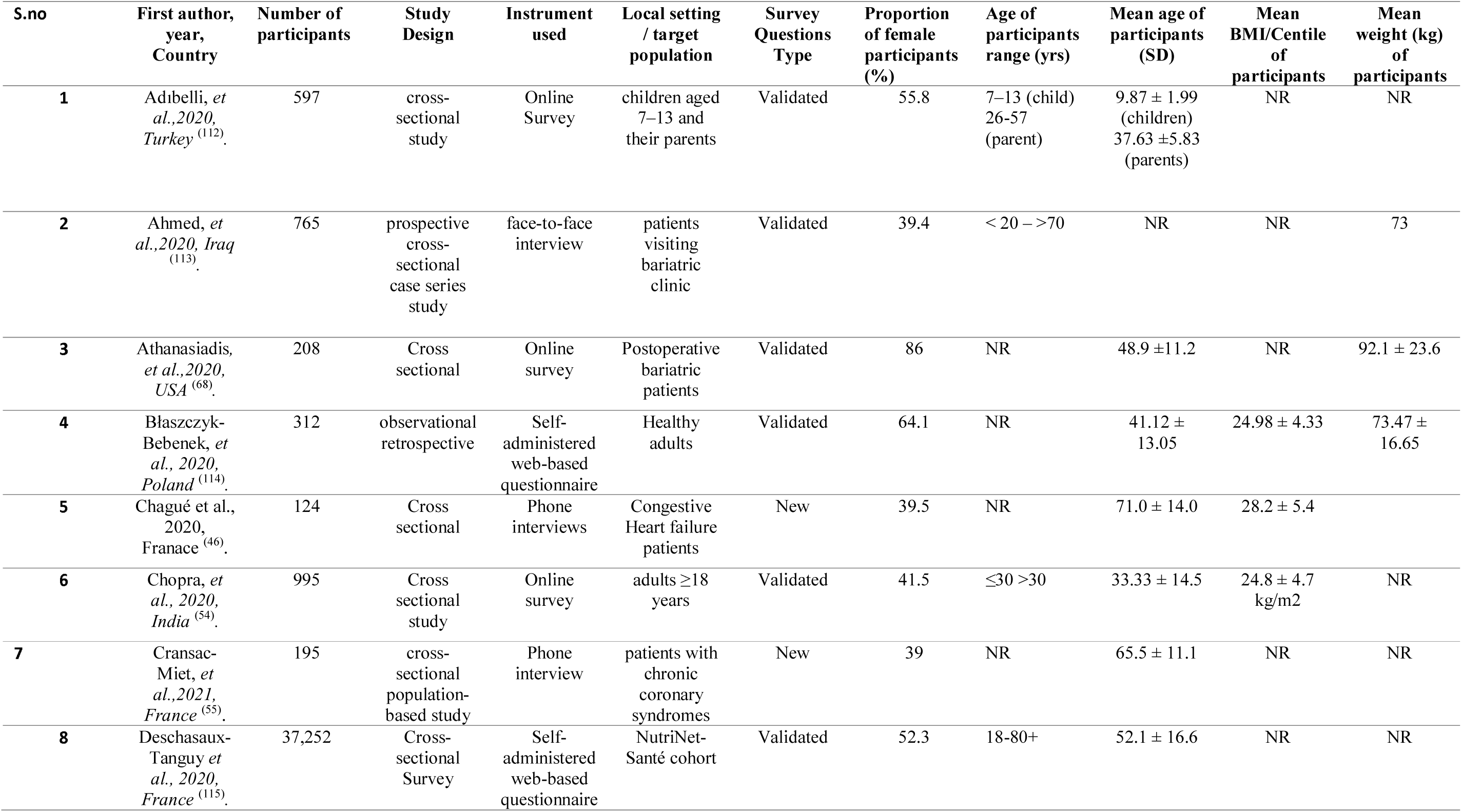
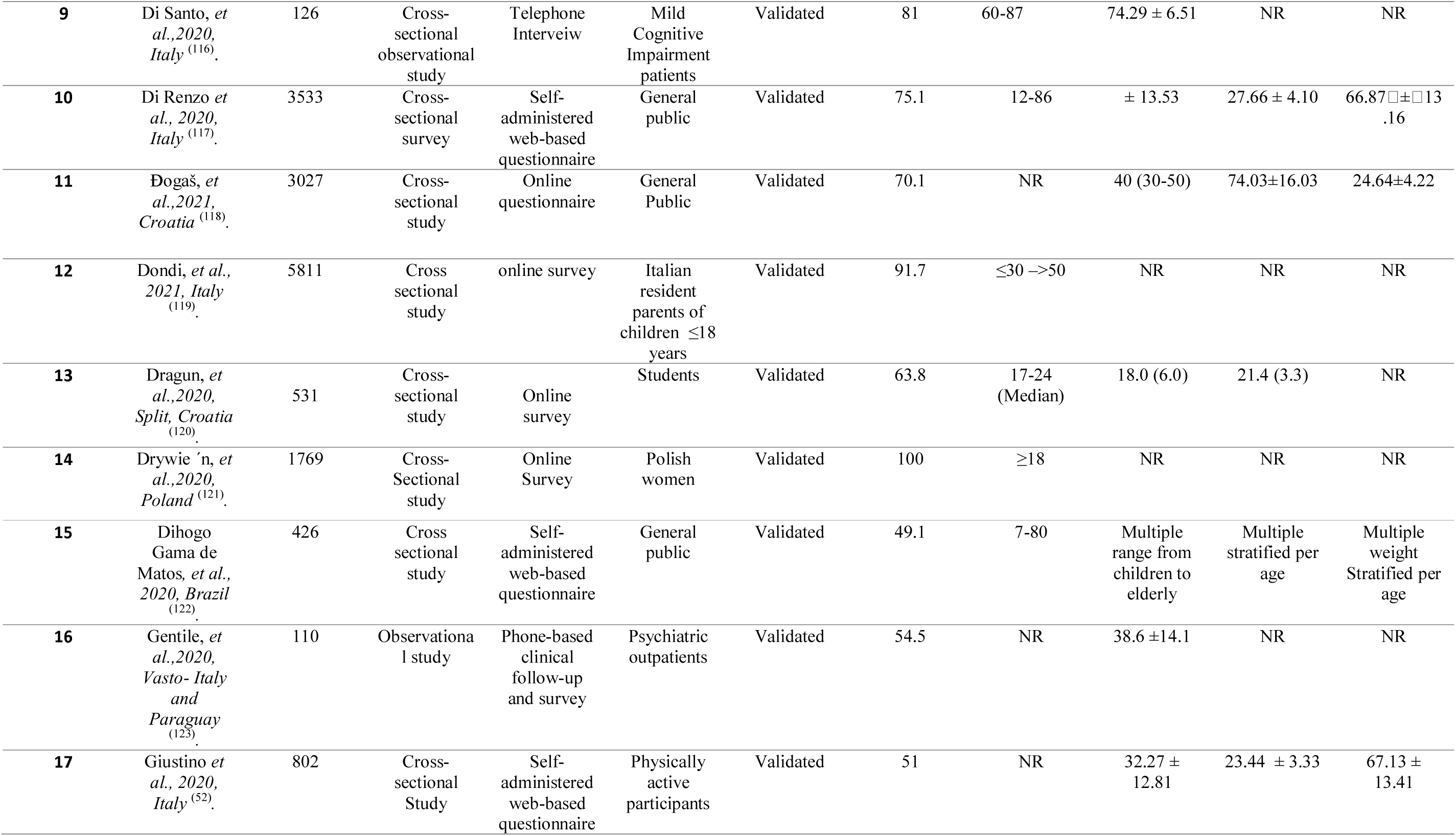
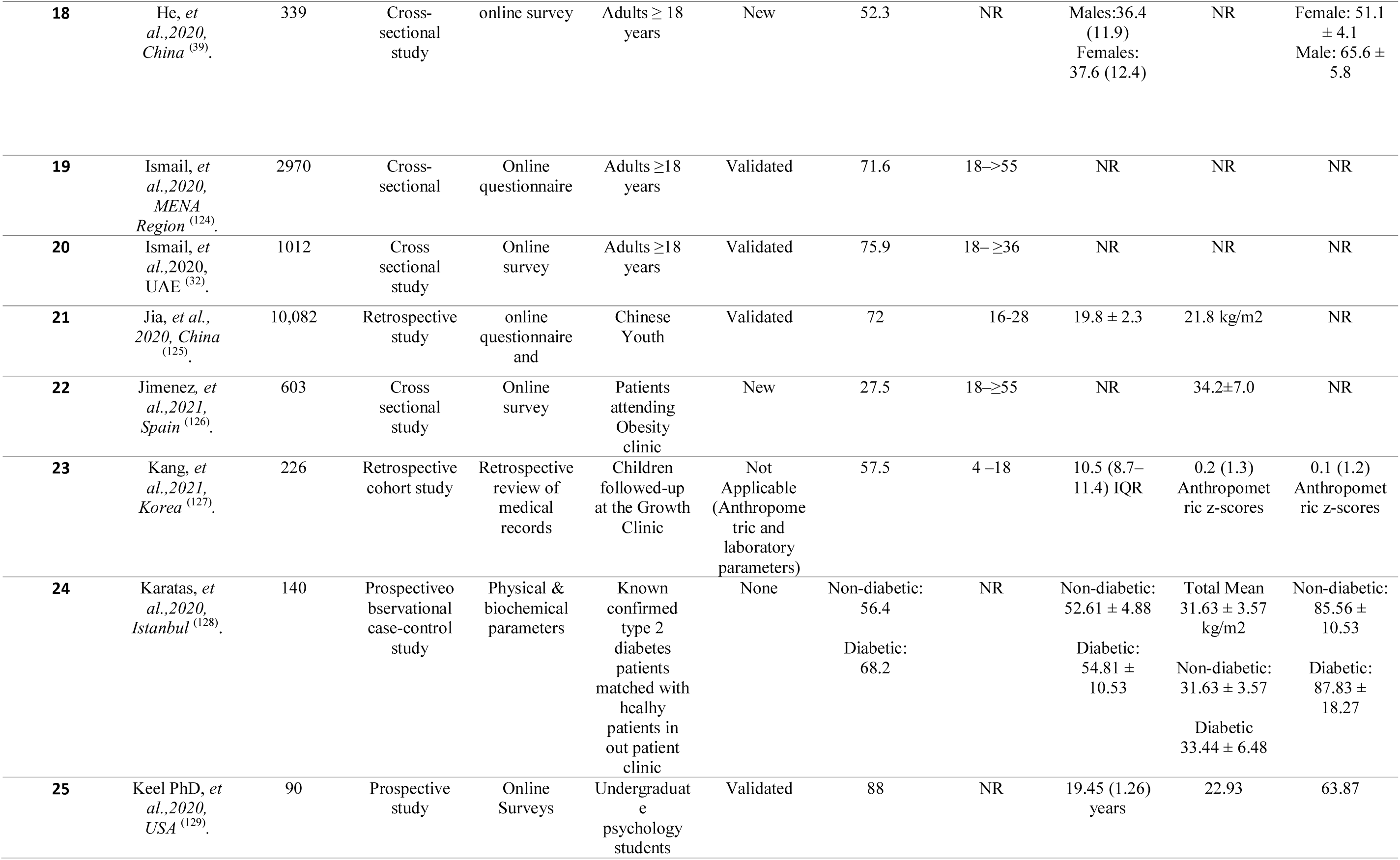
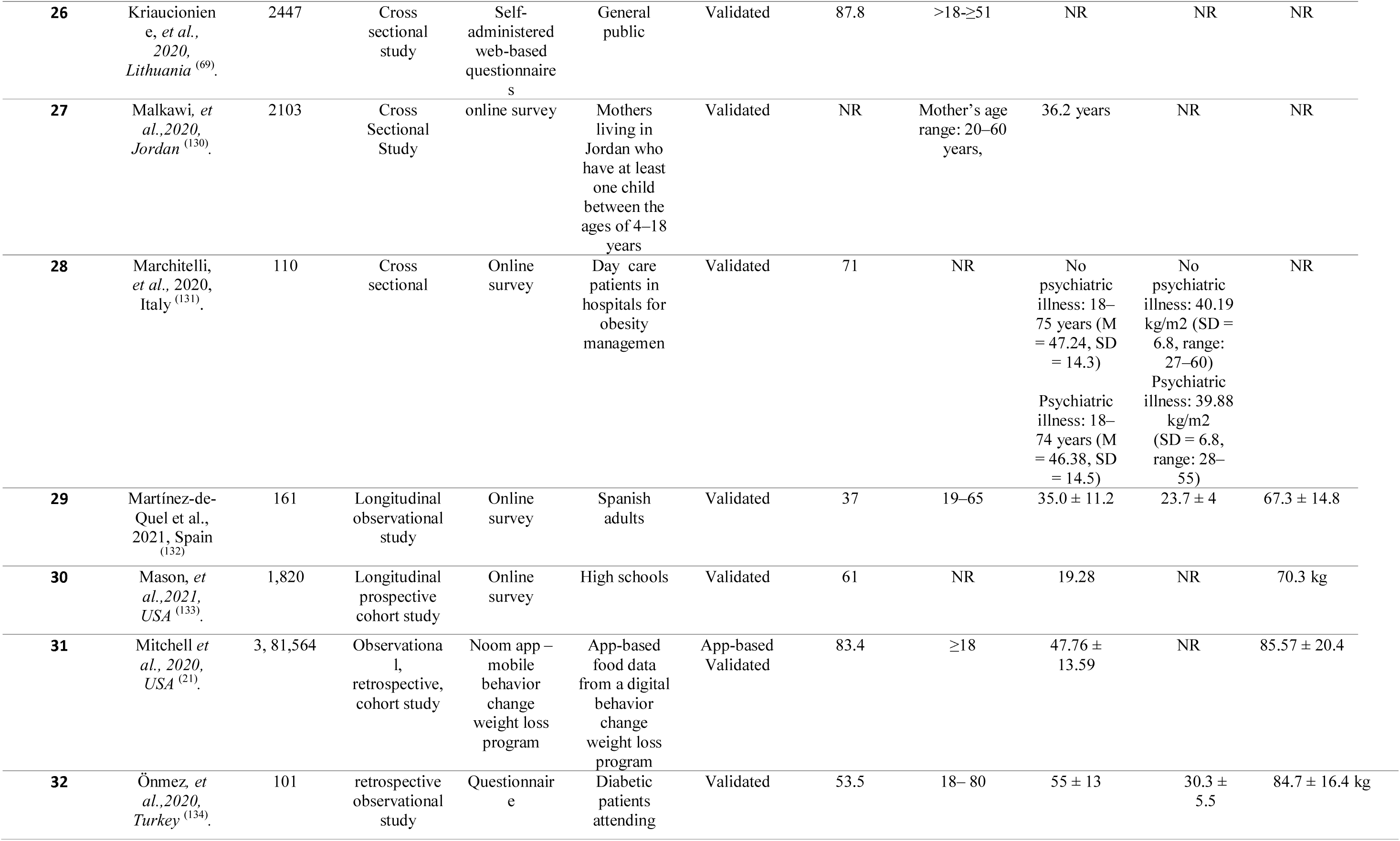
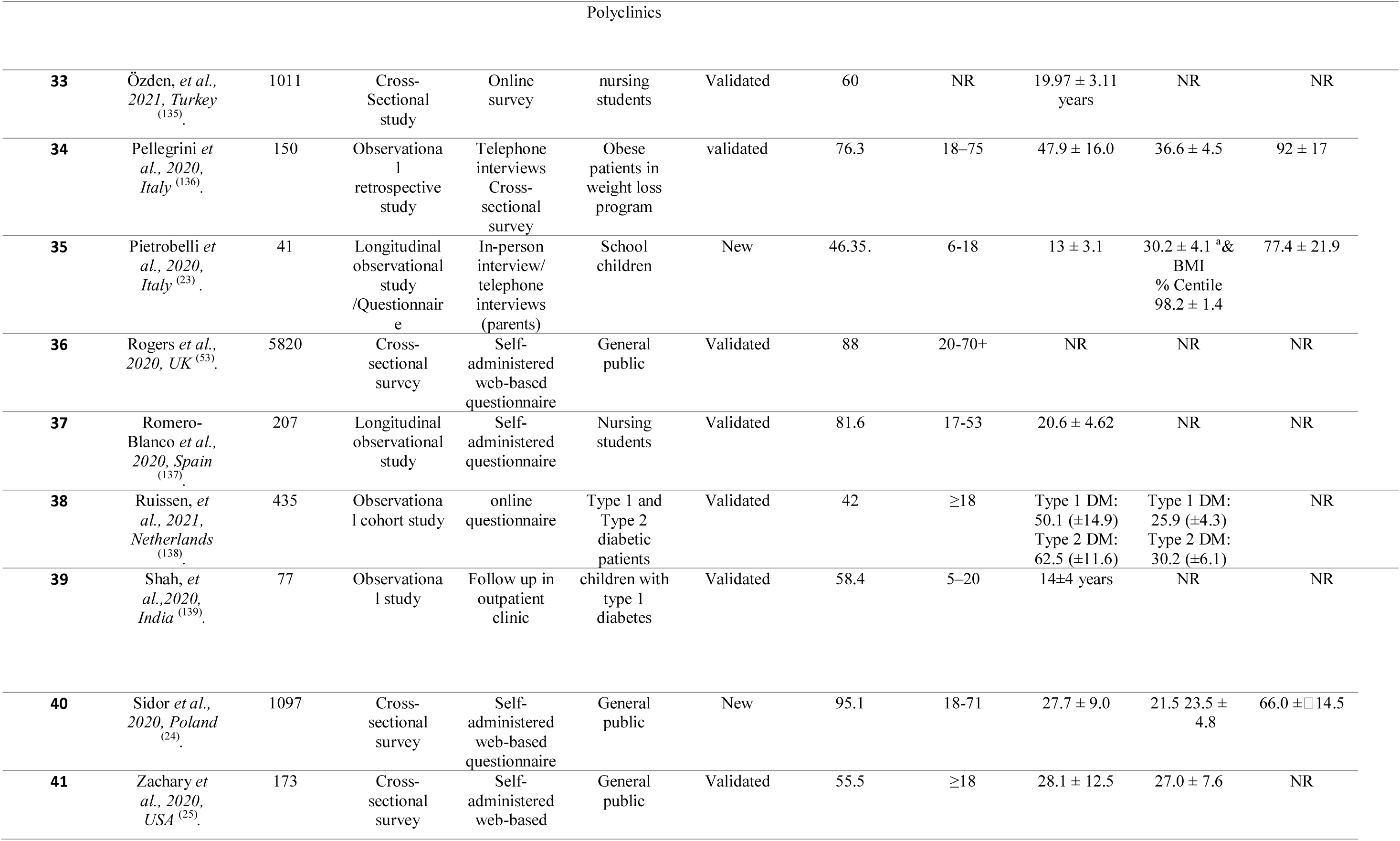
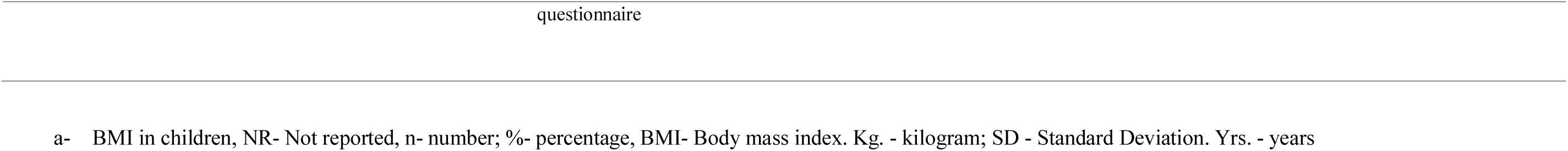
Included study characteristics.

The included studies had the following countries of origin: Brazil ^(59)^, China^(40,57)^, Croatia^(51,64)^, France ^(47,52,56)^, Jordan ^(58)^, India ^(48,55)^, Iraq ^(61)^, Italy^(21,23,24,35,36,45,45,53)^, Korea^(50)^, Lithuania ^(39)^, Netherlands ^(60)^, Poland ^(25,42,51)^, Spain^(44,46,63)^, Turkey ^(43,49,65,66)^, United Arab Emirates ^(33)^, United Kingdom ^(54)^ and the United States of America ^(26,38,41,67)^. Furthermore, multi-regional studies conducted intercontinentally ^(22)^, among eighteen countries in the Middle East and North Africa (MENA) region ^(34)^, and Paraguay and Italian based multinational research ^(62)^ are included in our analysis.

Altogether, the studies enrolled 469,362 participants. The participants ranged in age from 6 to 86 years, and the mean ages for the individuals studied ranged from 9.9 to 74.3 years. The proportion of female participants ranged from 37% to 100%. The number of participants in the included studies ranged from 41 to 381,564. All studies included both male and female participants except one study ^(37)^. The duration of confinement for the selected studies for this systematic review ranged between 1 and 24 weeks.

### Impact of confinement on body weight

In our study, 7.2%–72.4% of all participants including both adults and children, experienced an increase in body weight during the confinement periods ^(21,23,25,26,33,35–37,39–41,43,44,44–49,49–52,55,57,57–62,64–68)^(Figure 2). The mean weight gain ranged from 0.6 (±1.3) to 3.0 (±2.4) kg. There was a higher weight gain among participants who self-reported stress^(26,45,55,56,58,61,62)^, anxiety and depression ^(23,52,58,61,62)^. Weight loss was observed in 11.1%–32.0% of participants^(21,25,33,35,37,40,51,52,55,60,65,68)^. The mean experienced weight loss ranged from 2.0 (±1.4) to 2.9 (±1.5) kg.

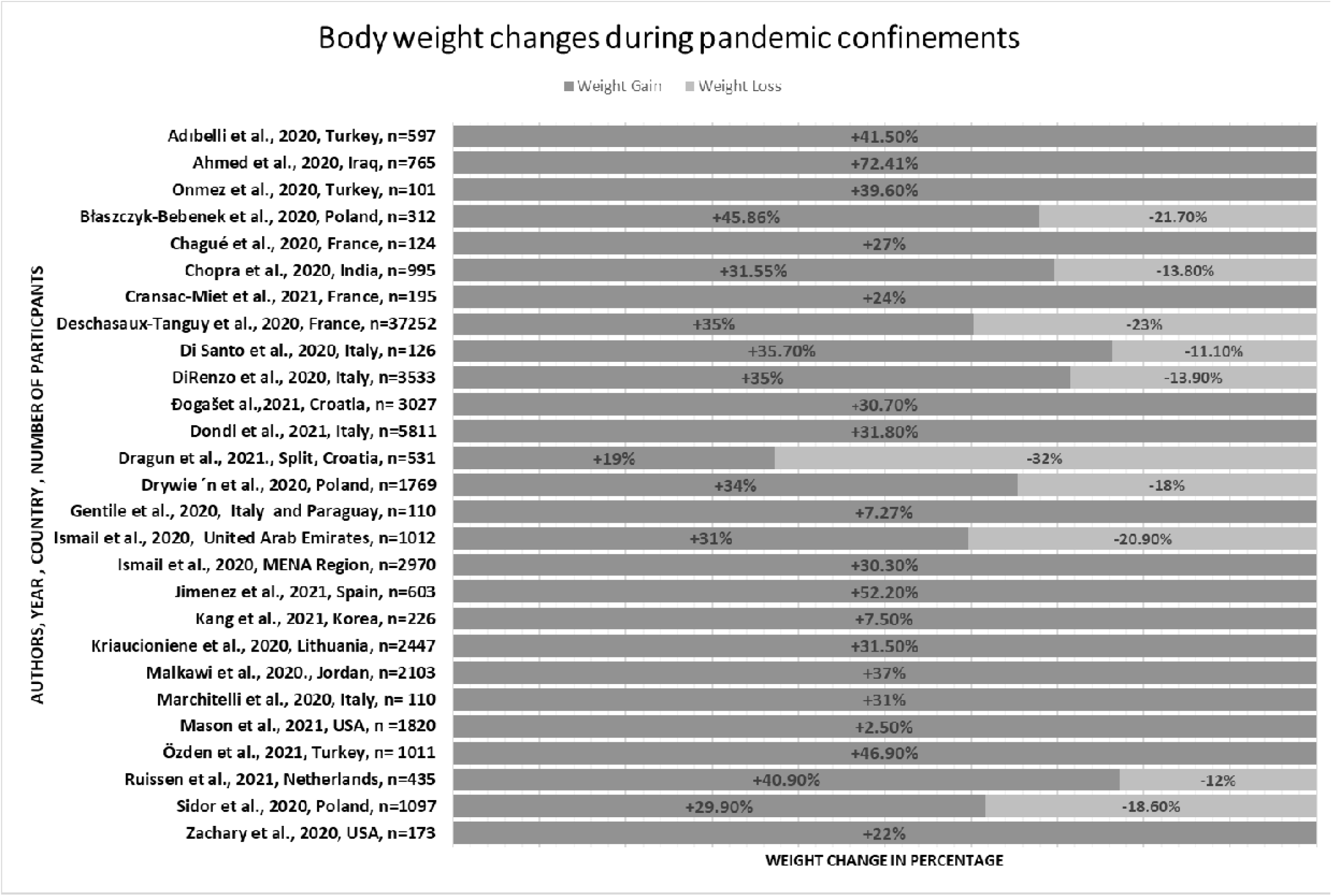

### Dietary and other lifestyle behavior changes during confinement

Table 3 describes dietary and behavioral changes that were caused by pandemic-related confinements. Most studies reported an increase in food intake associated with increased snacking ^(21,23,25,26,33,34,36,36,37,39,41,43,44,51,52,55,68)^ and all these studies documenting perceived weight gain ^(21,23,25,26,33,34,36,37,39,41,43,44,51,52,55,68)^.

**Table 3:**
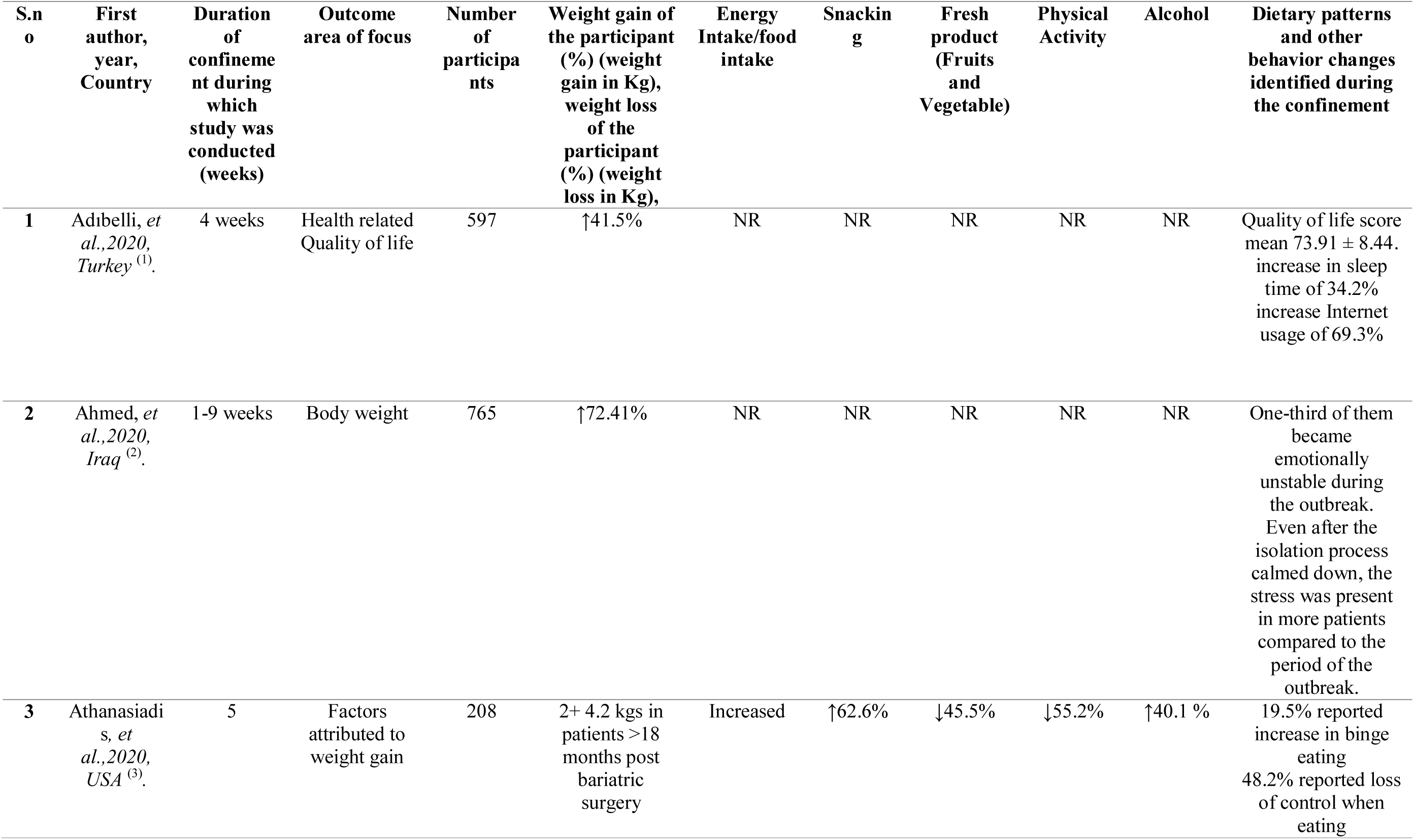
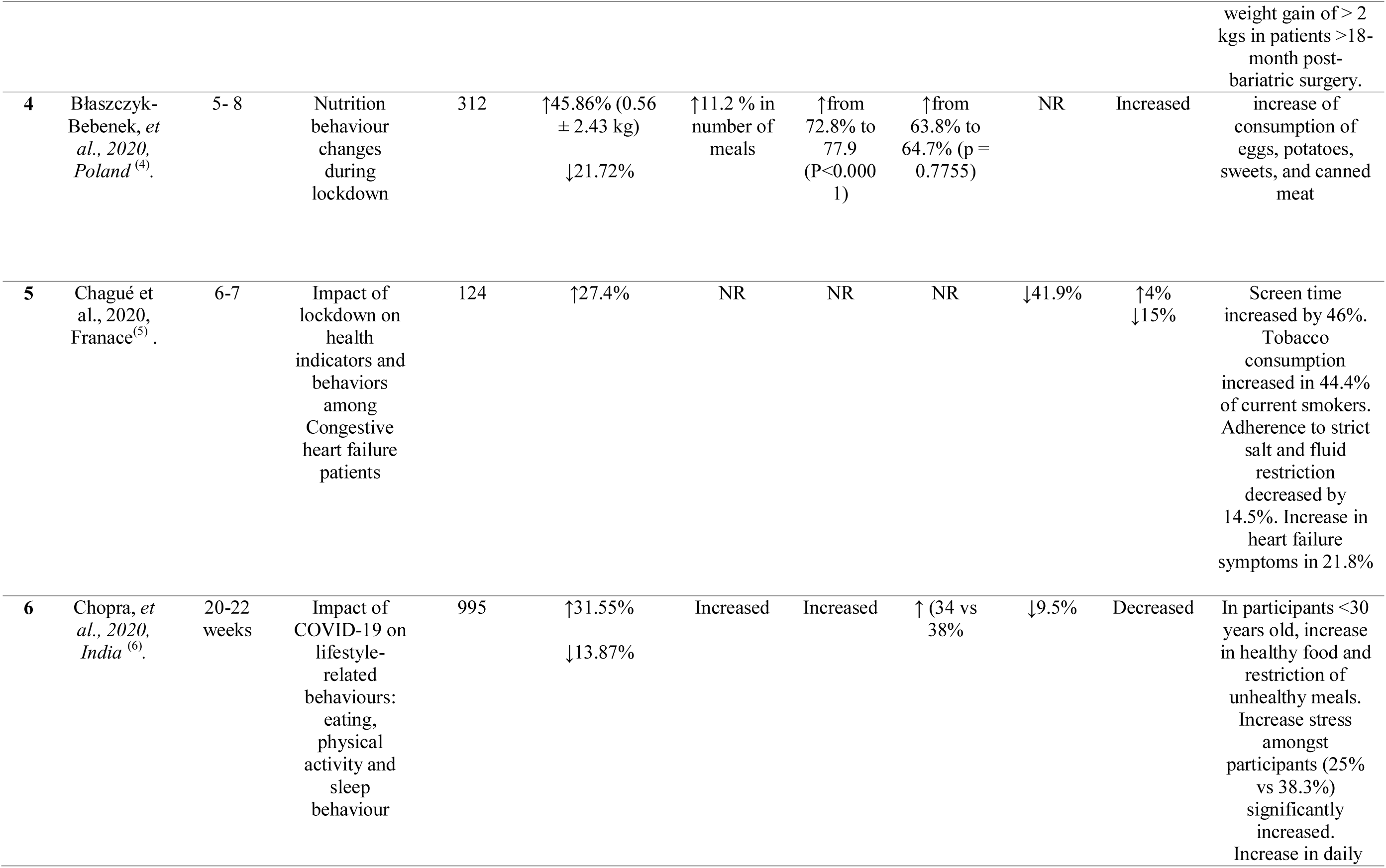
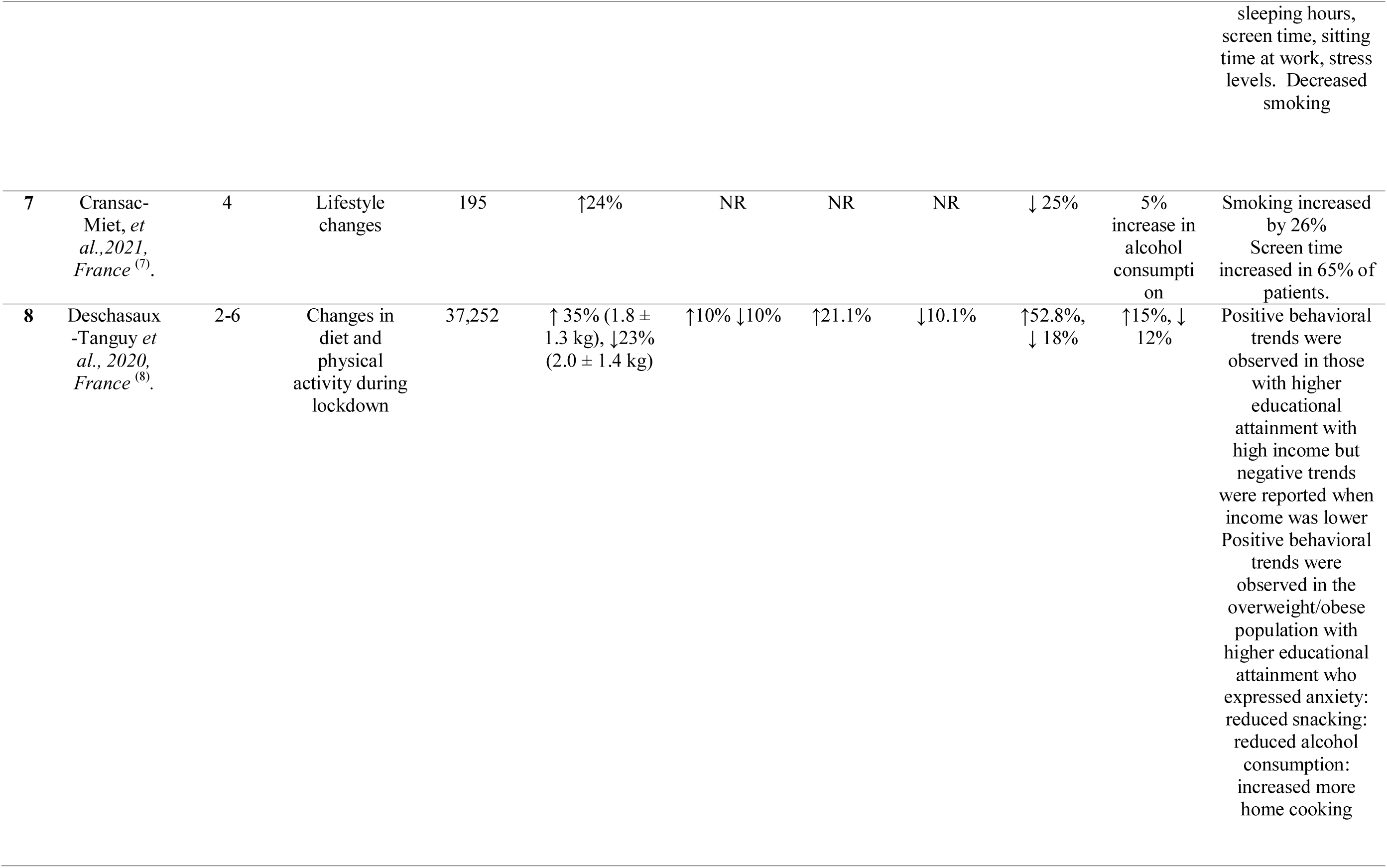
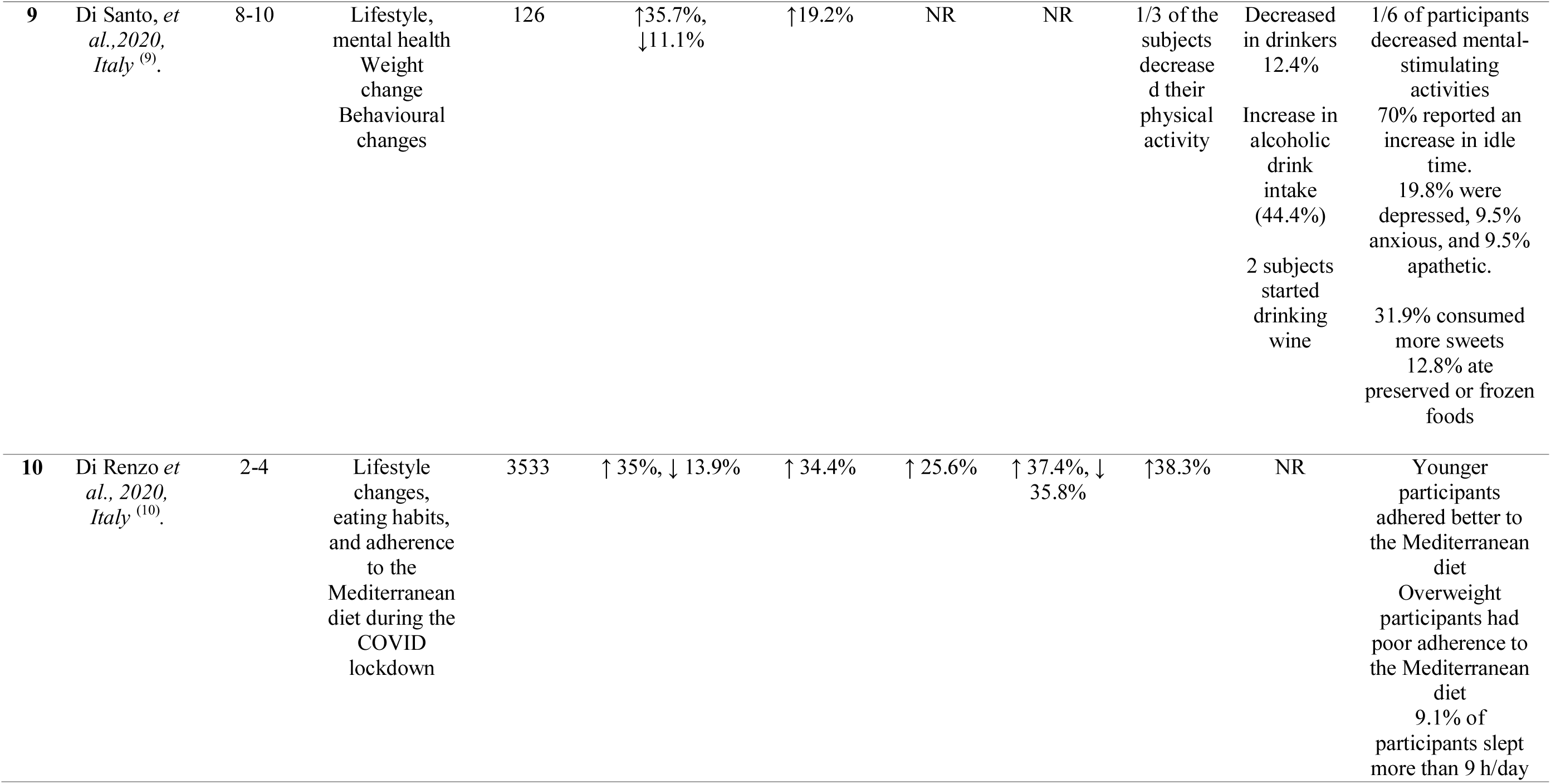
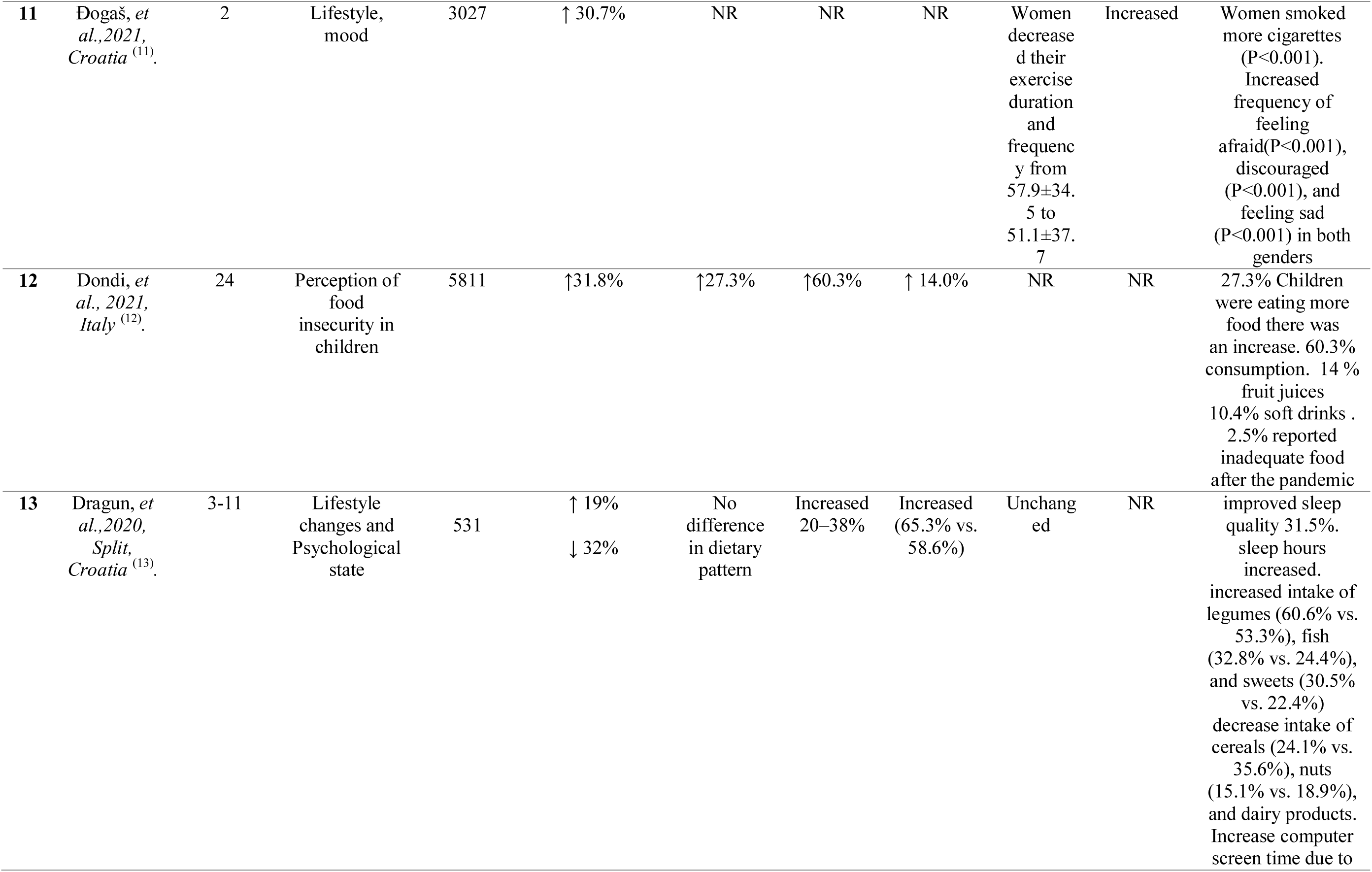
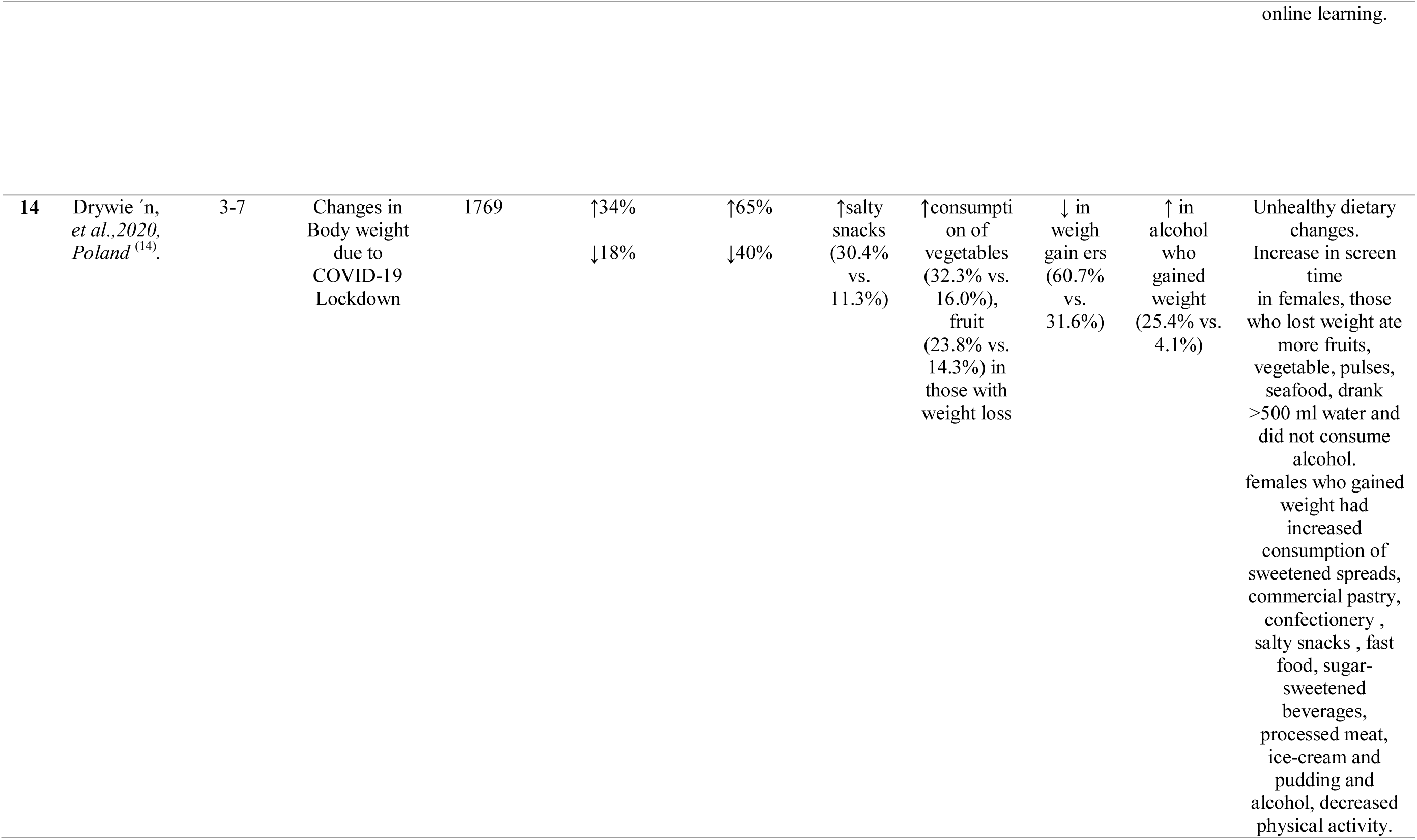
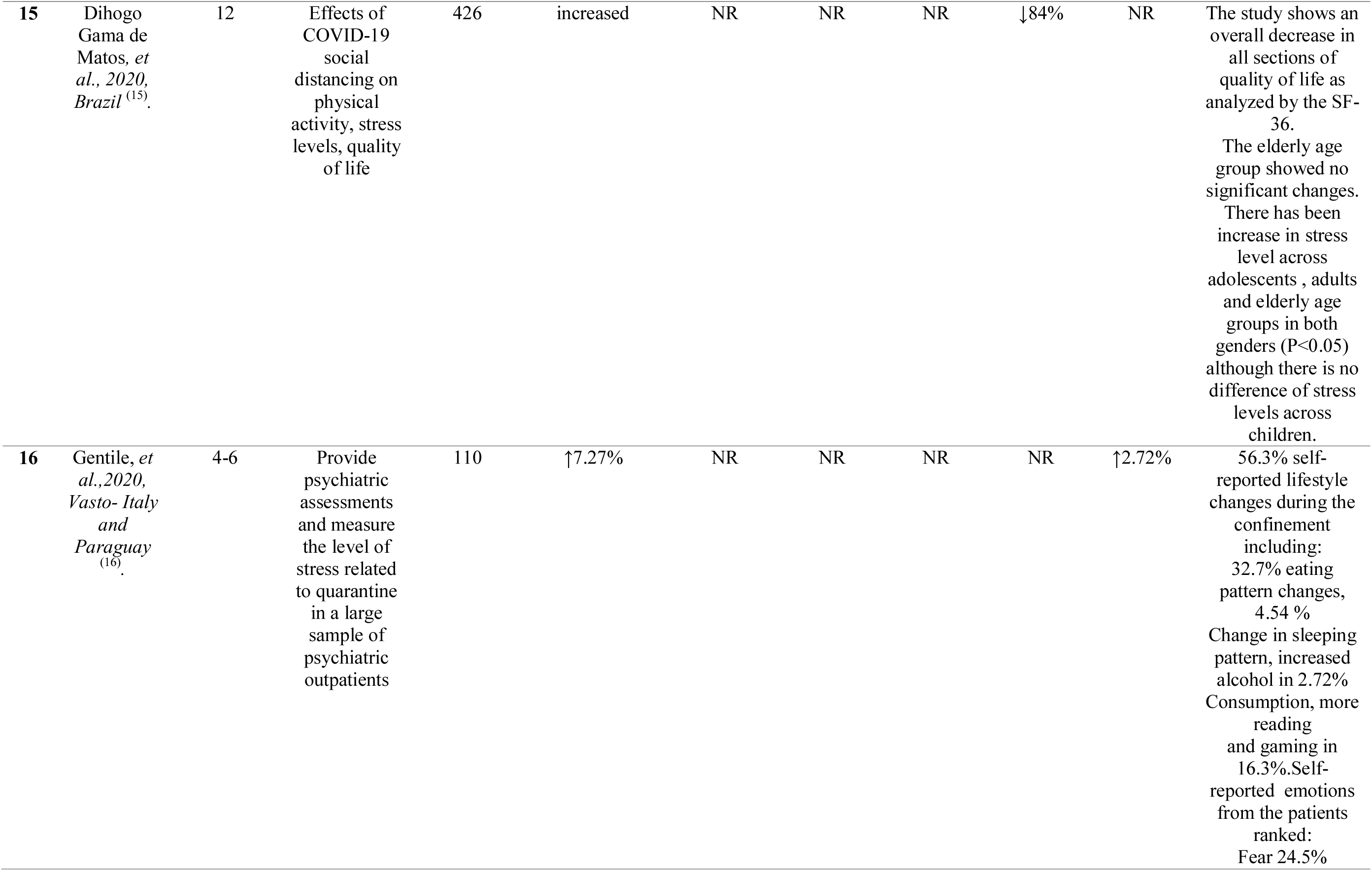
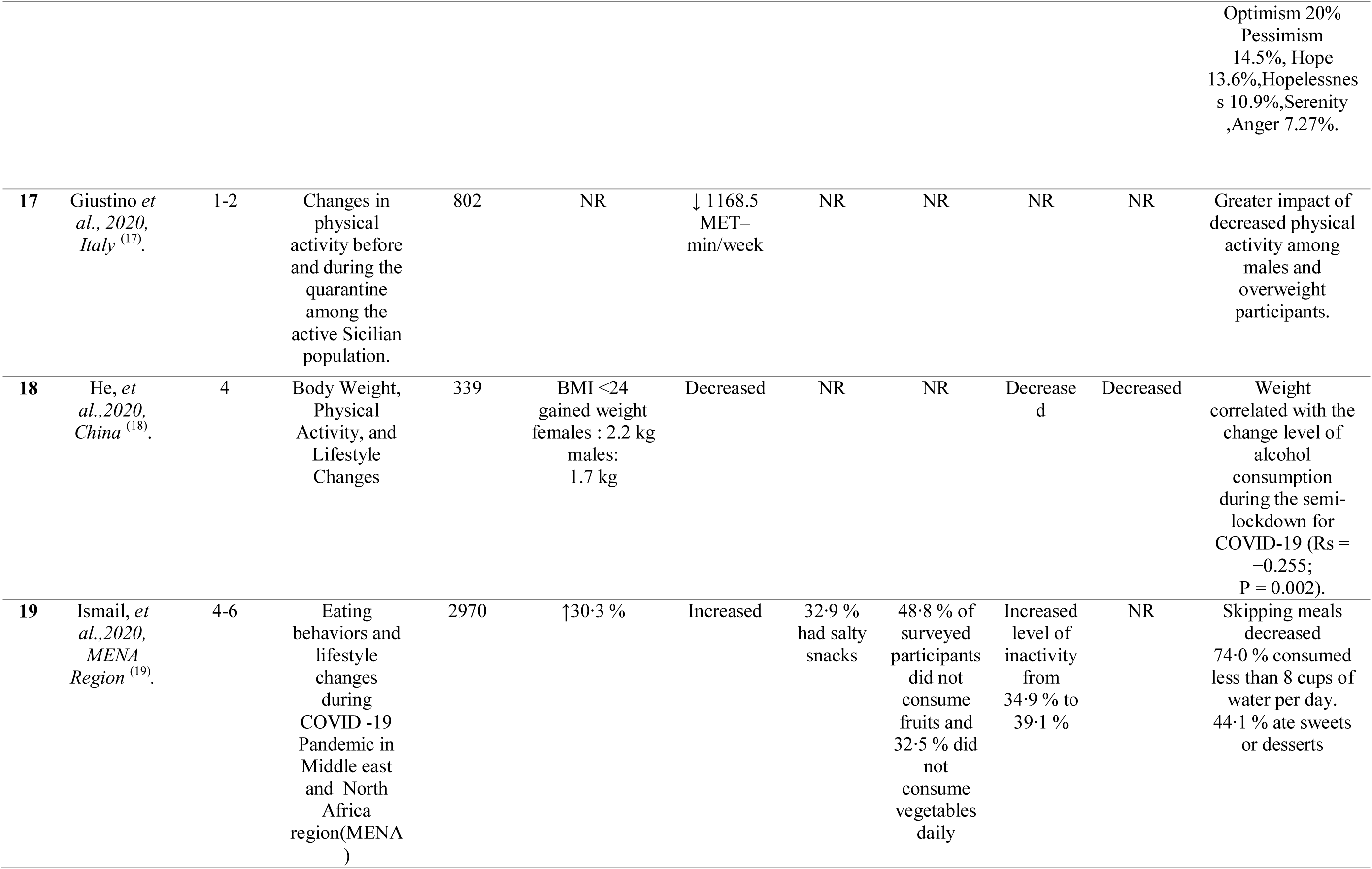
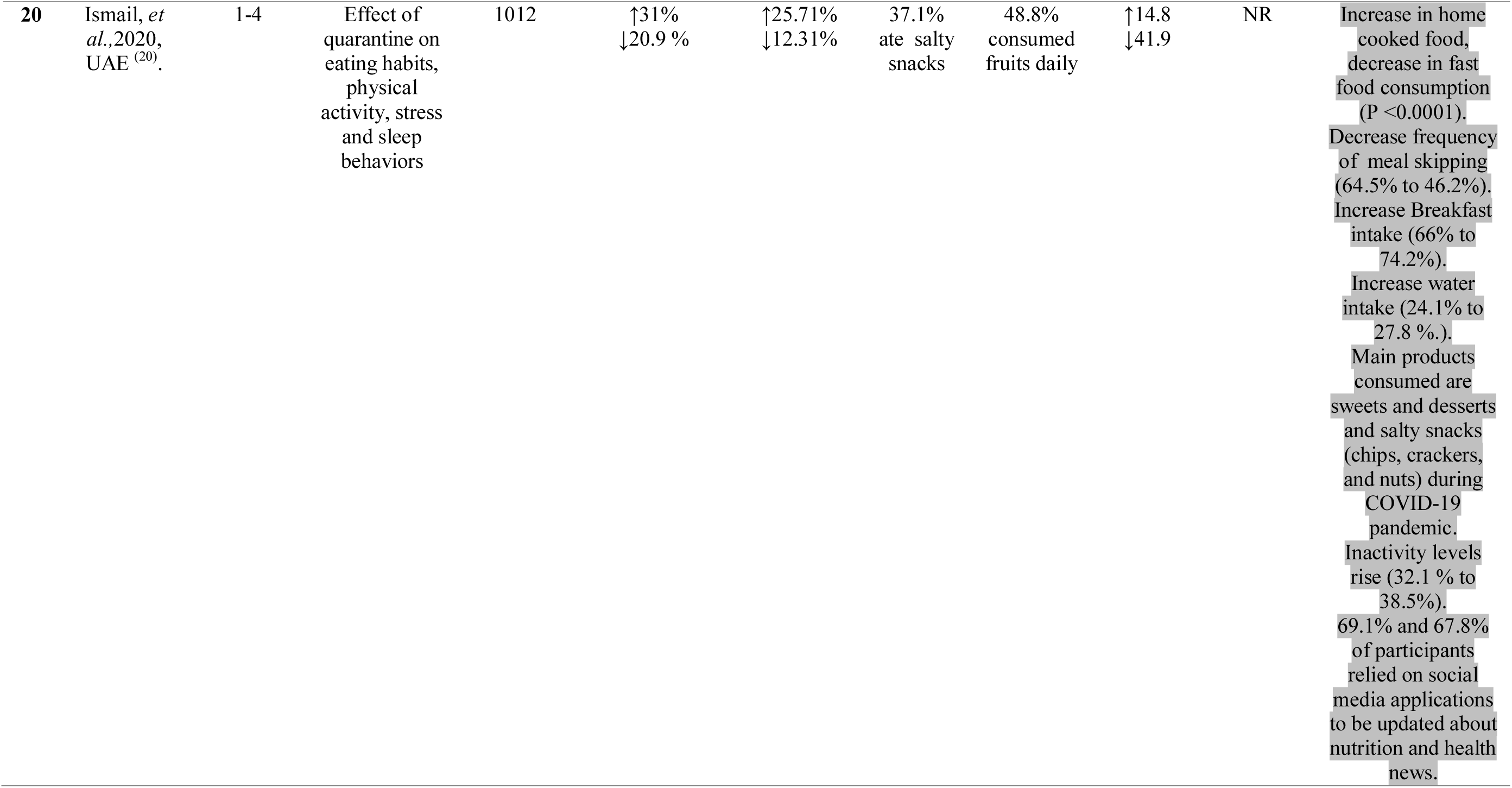
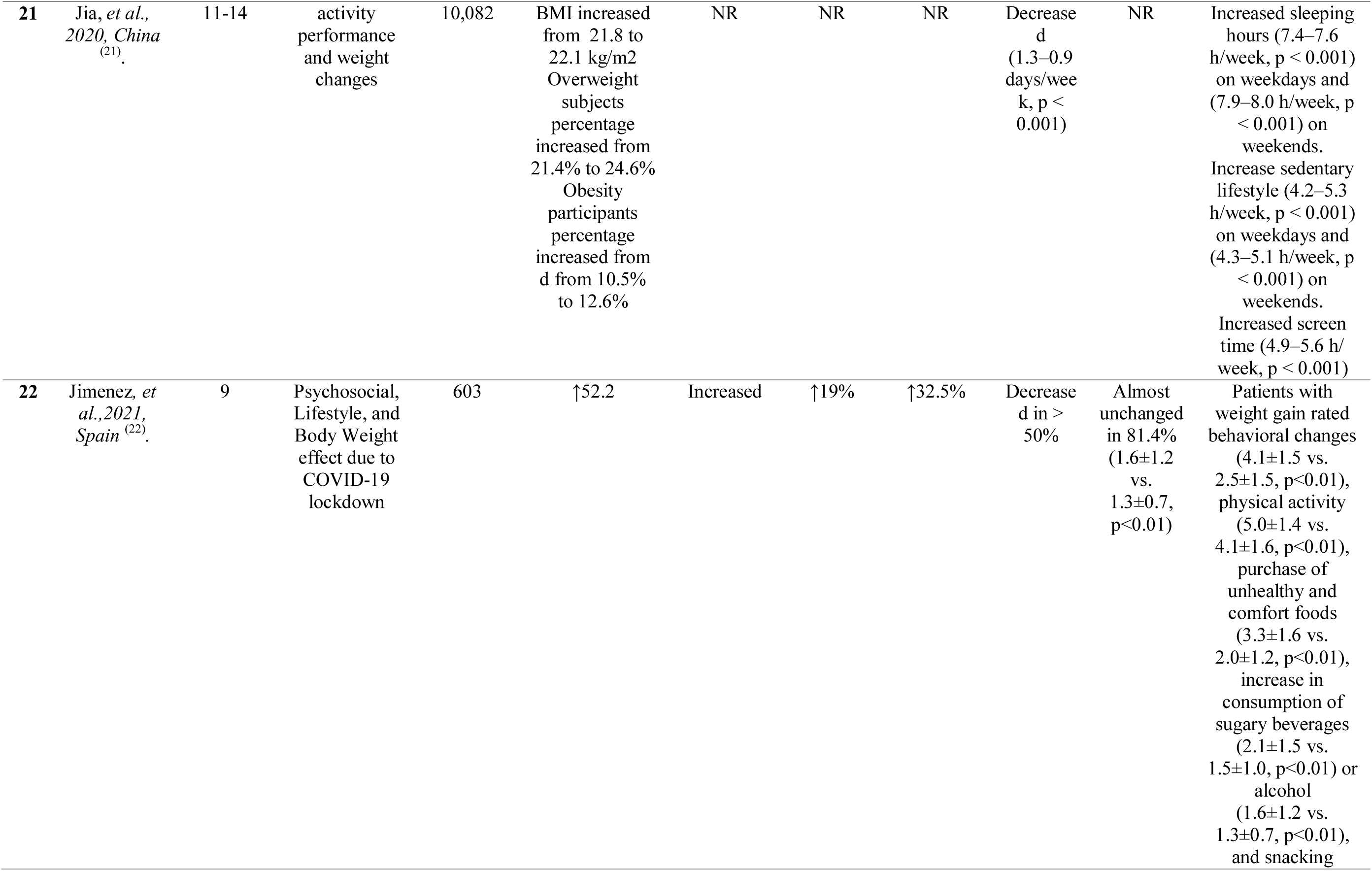
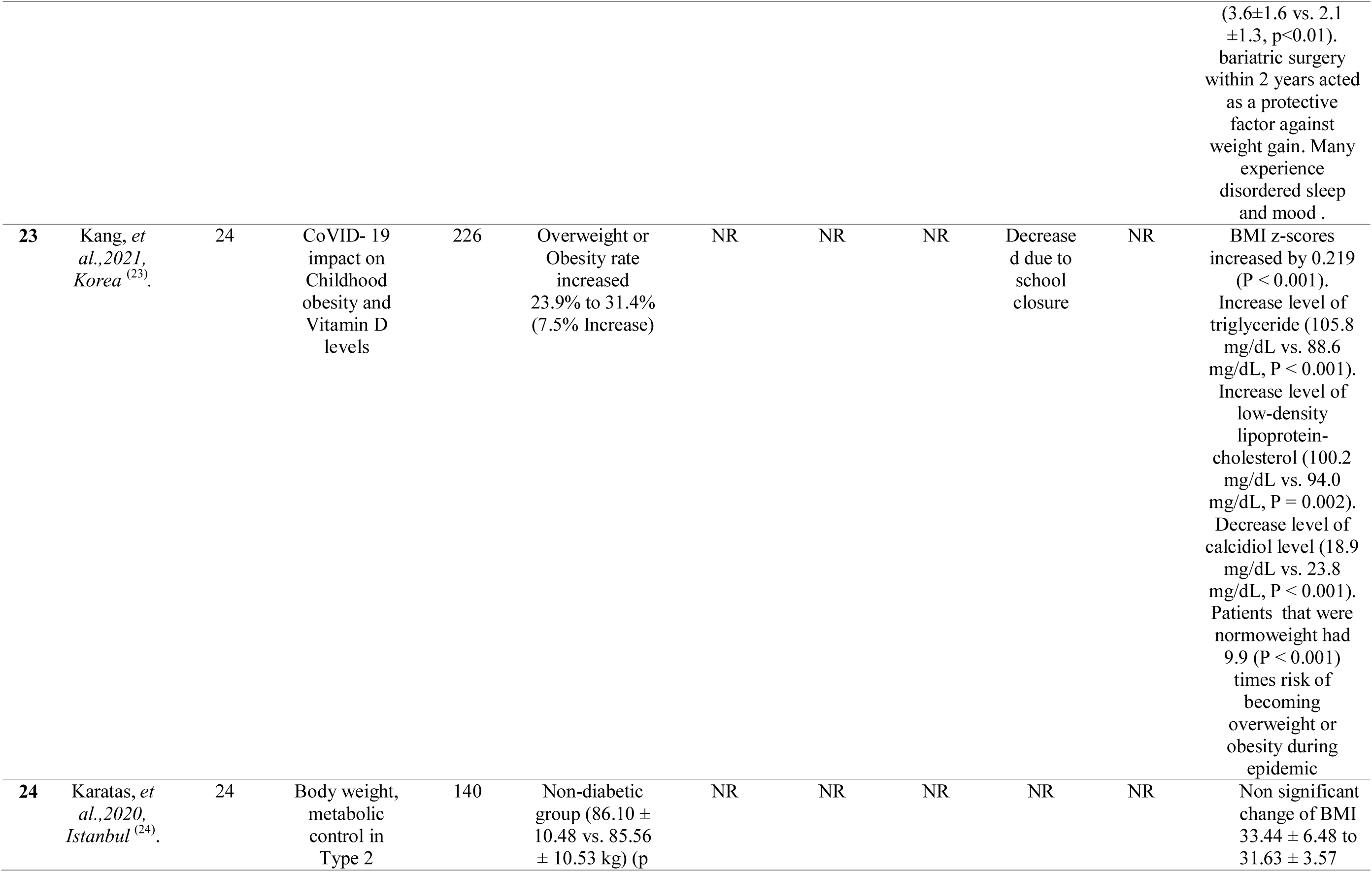
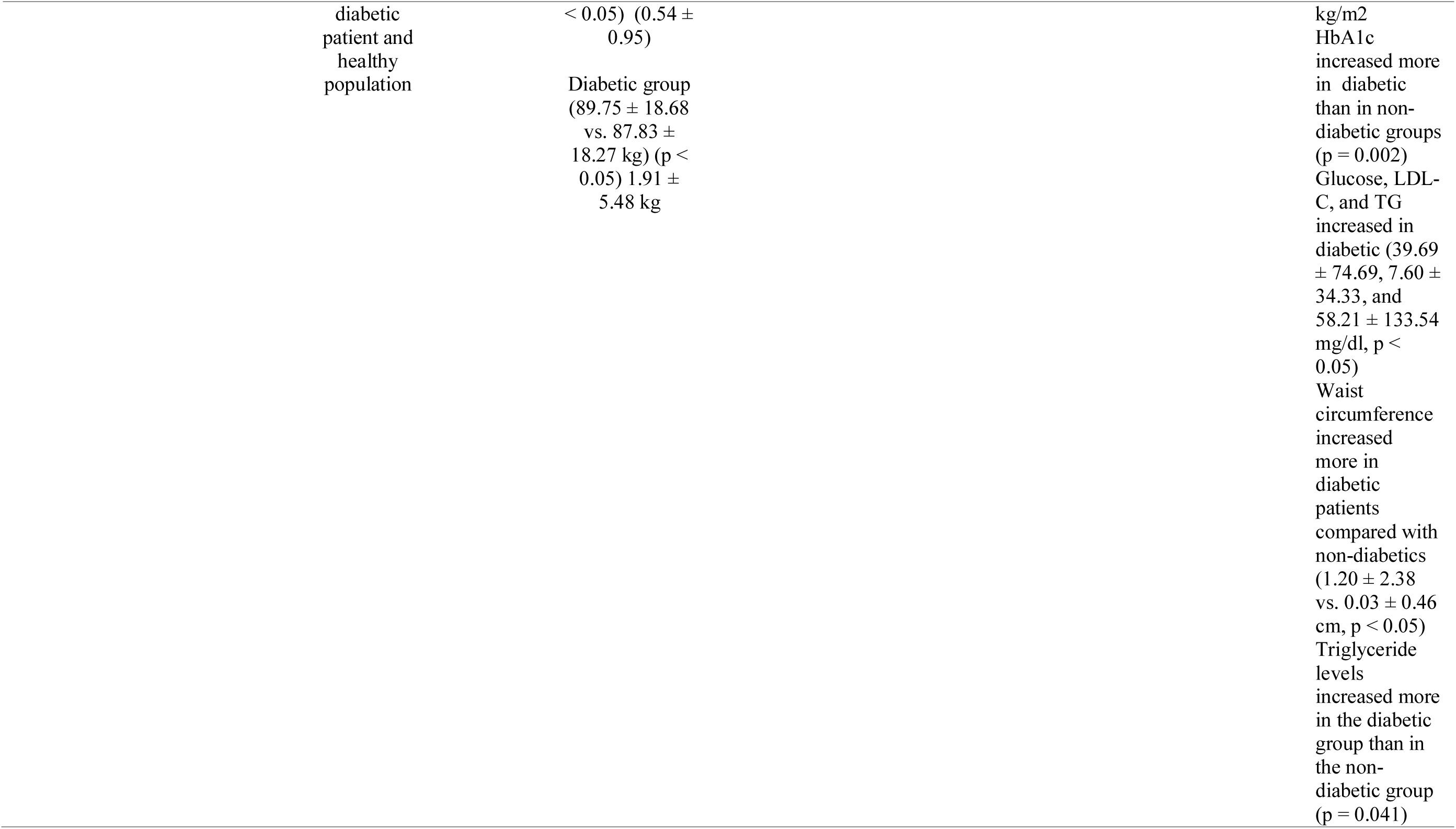
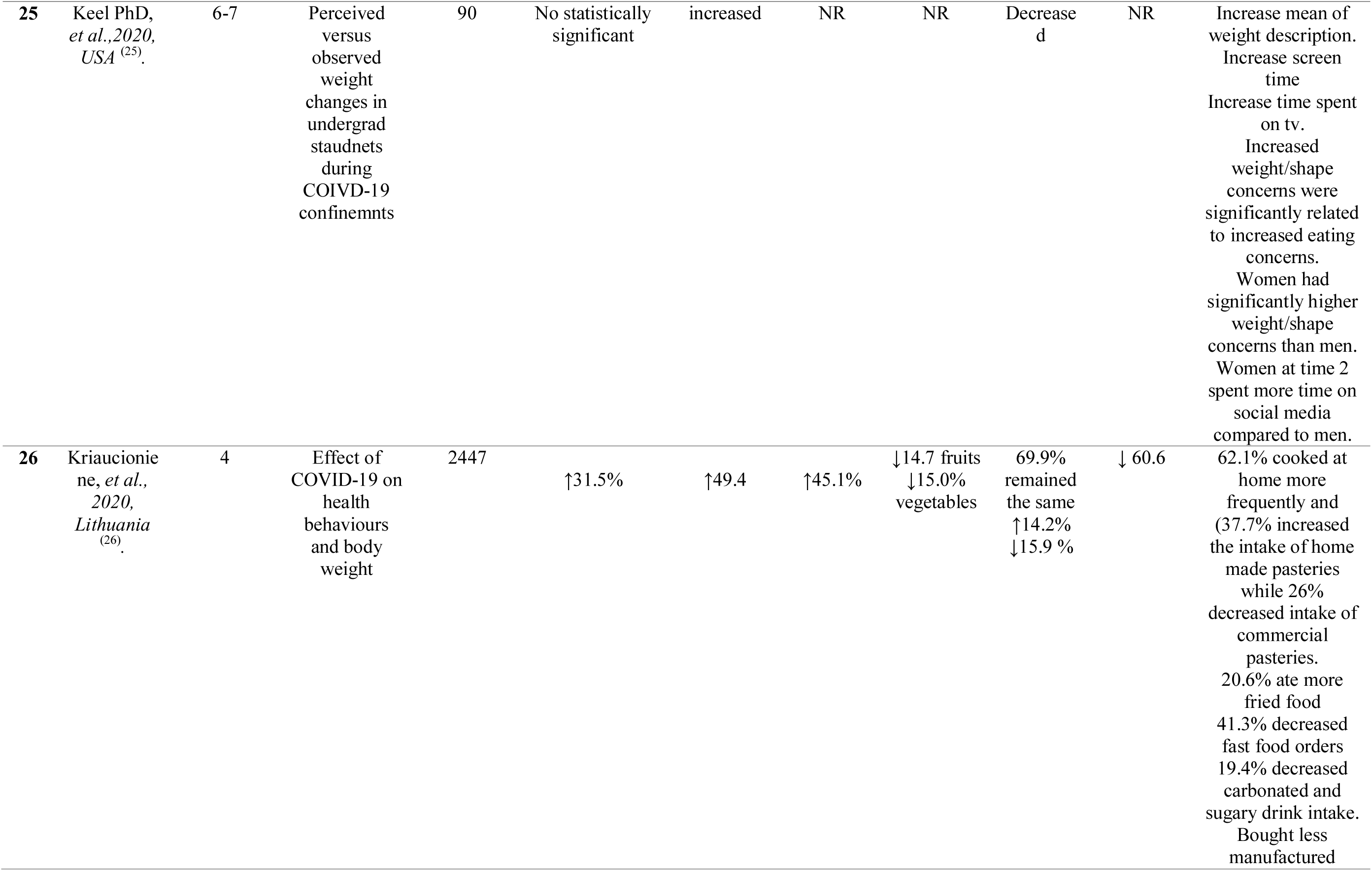
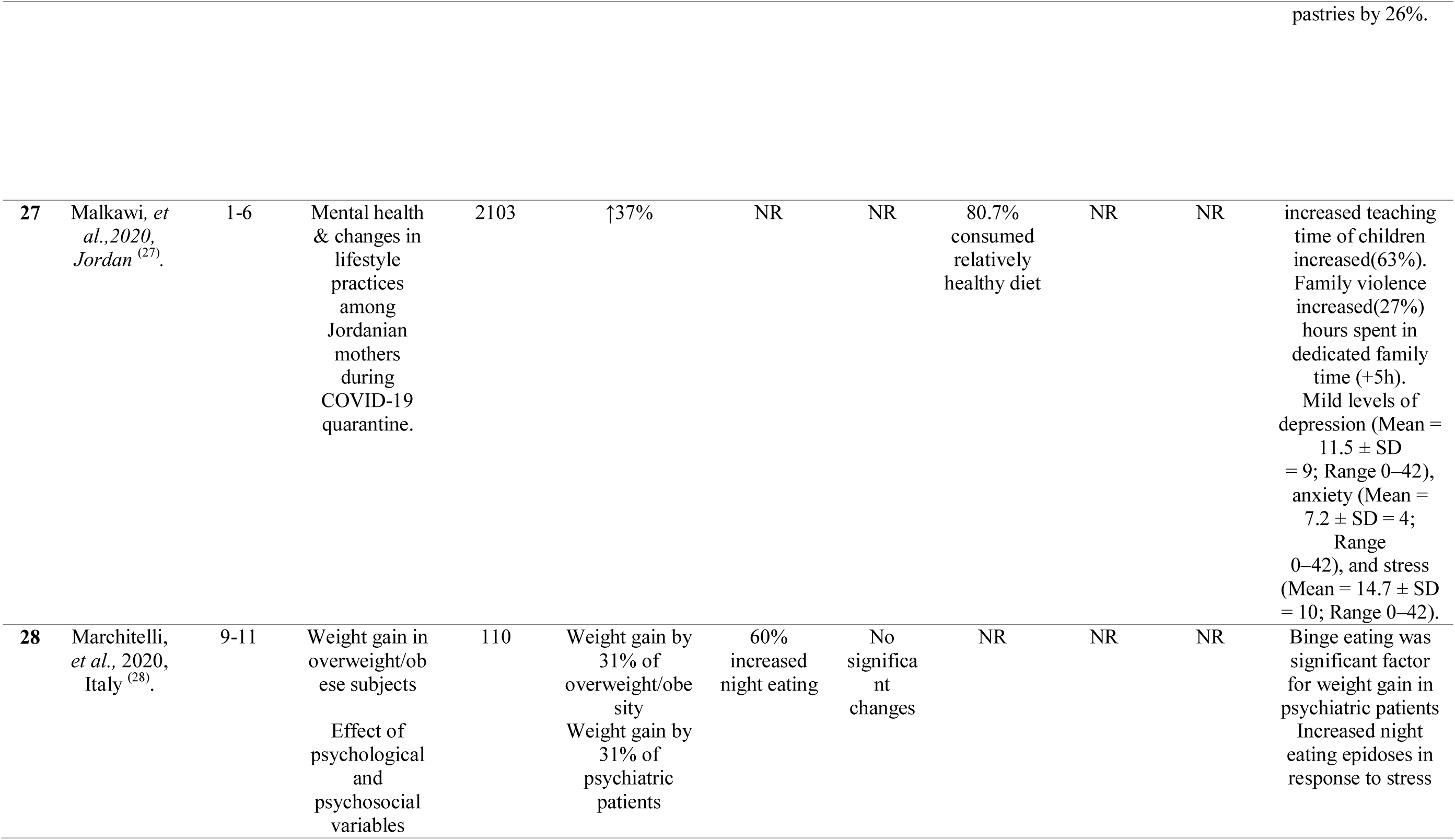
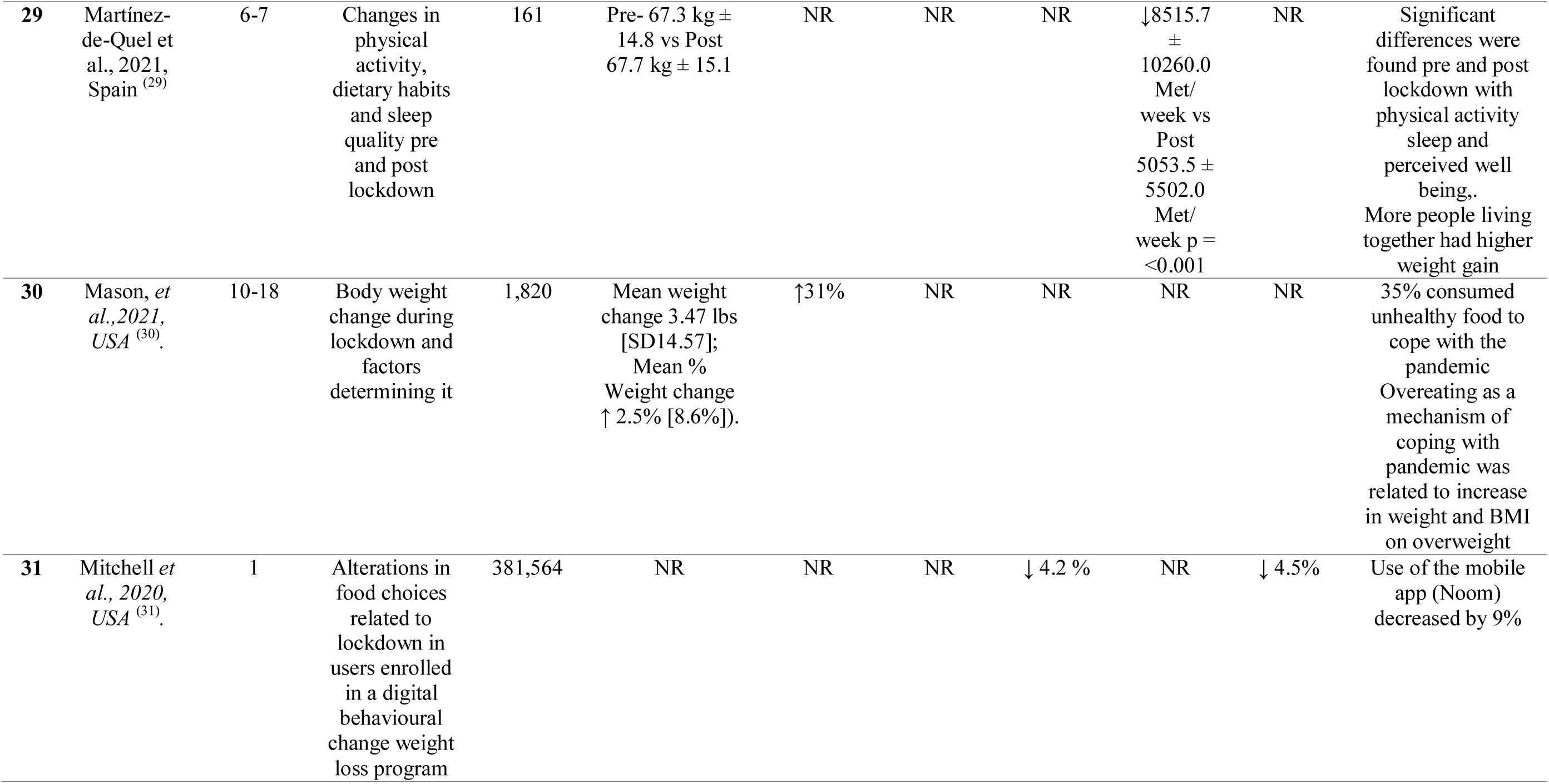
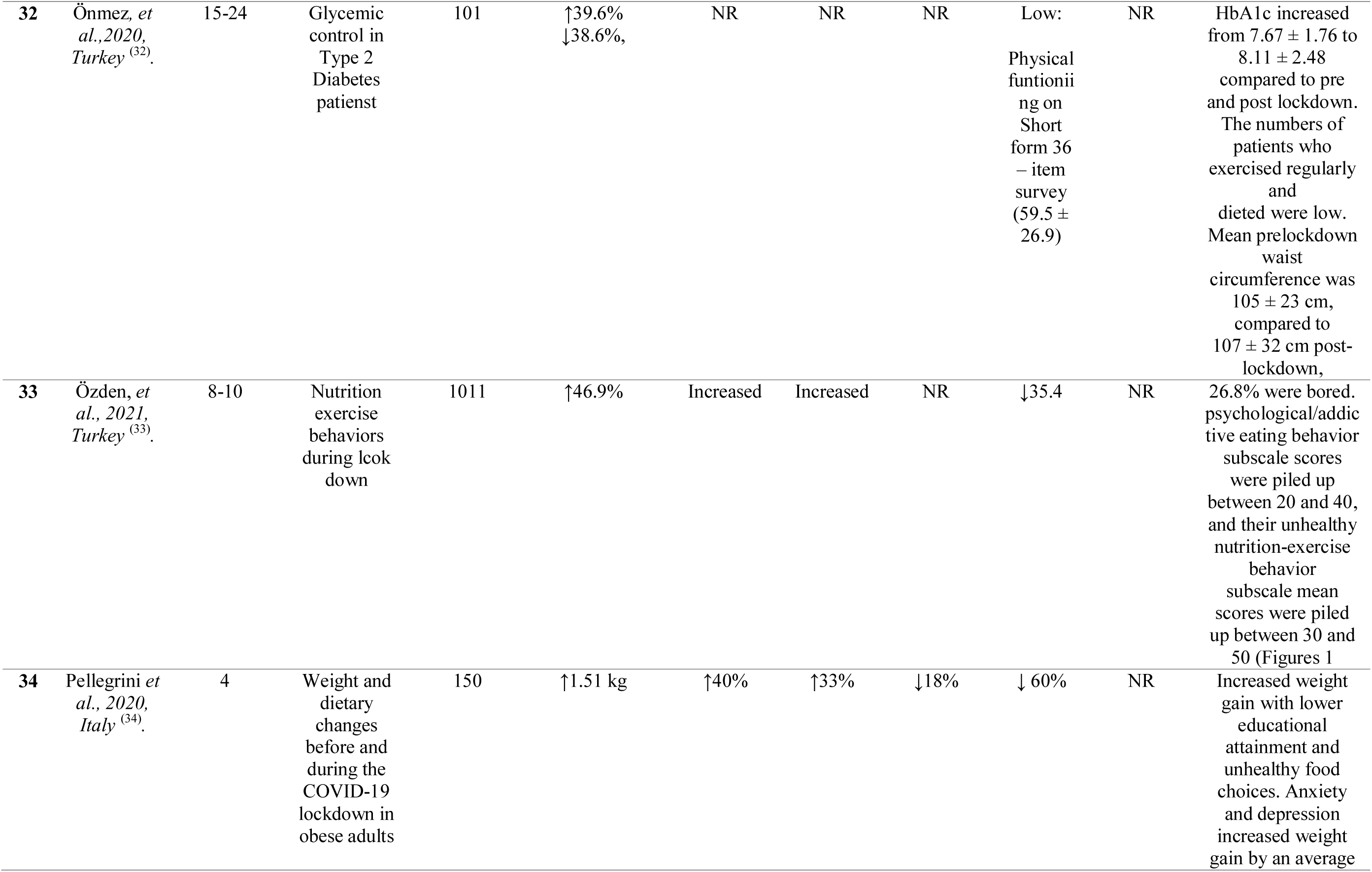
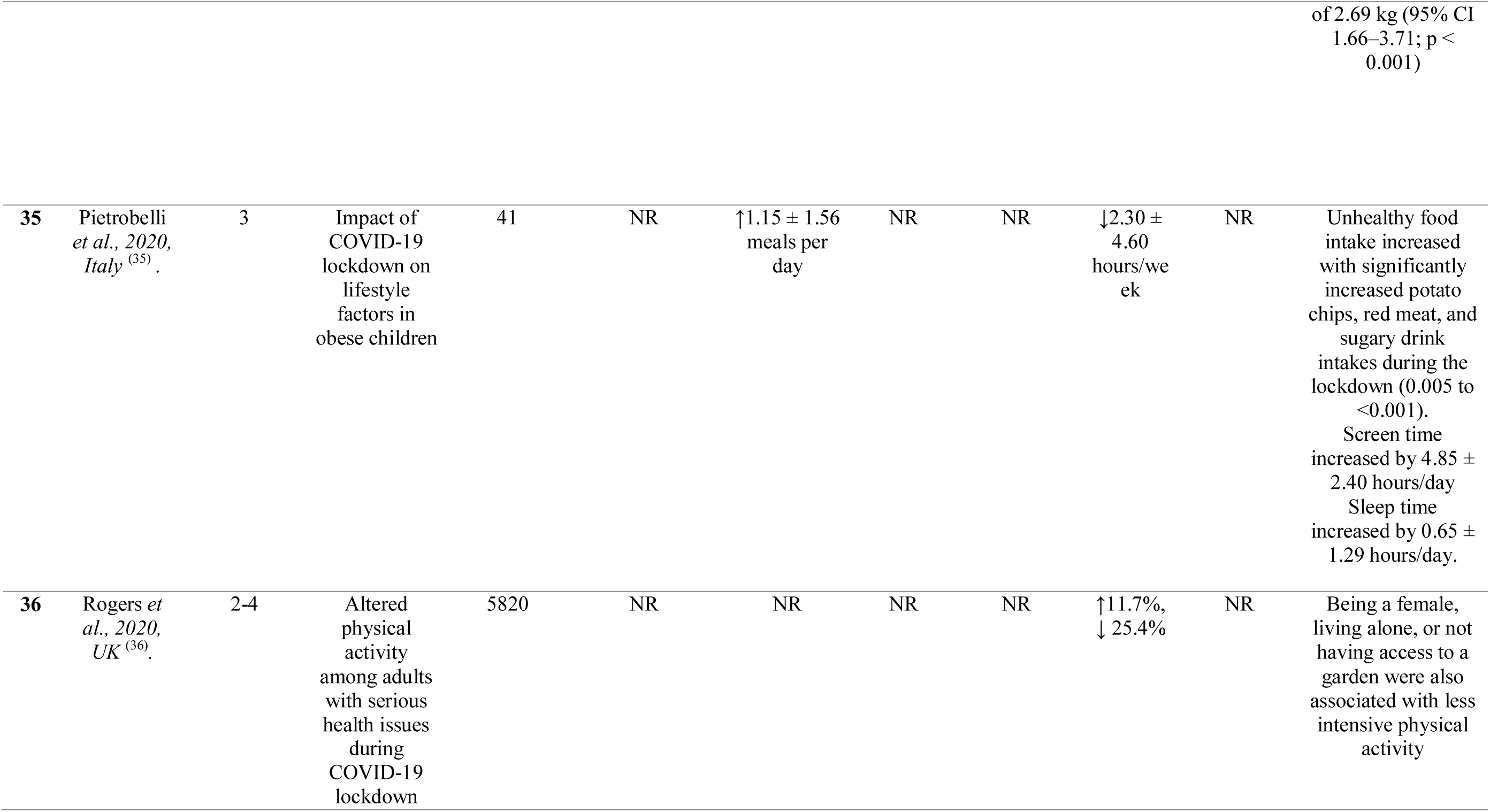
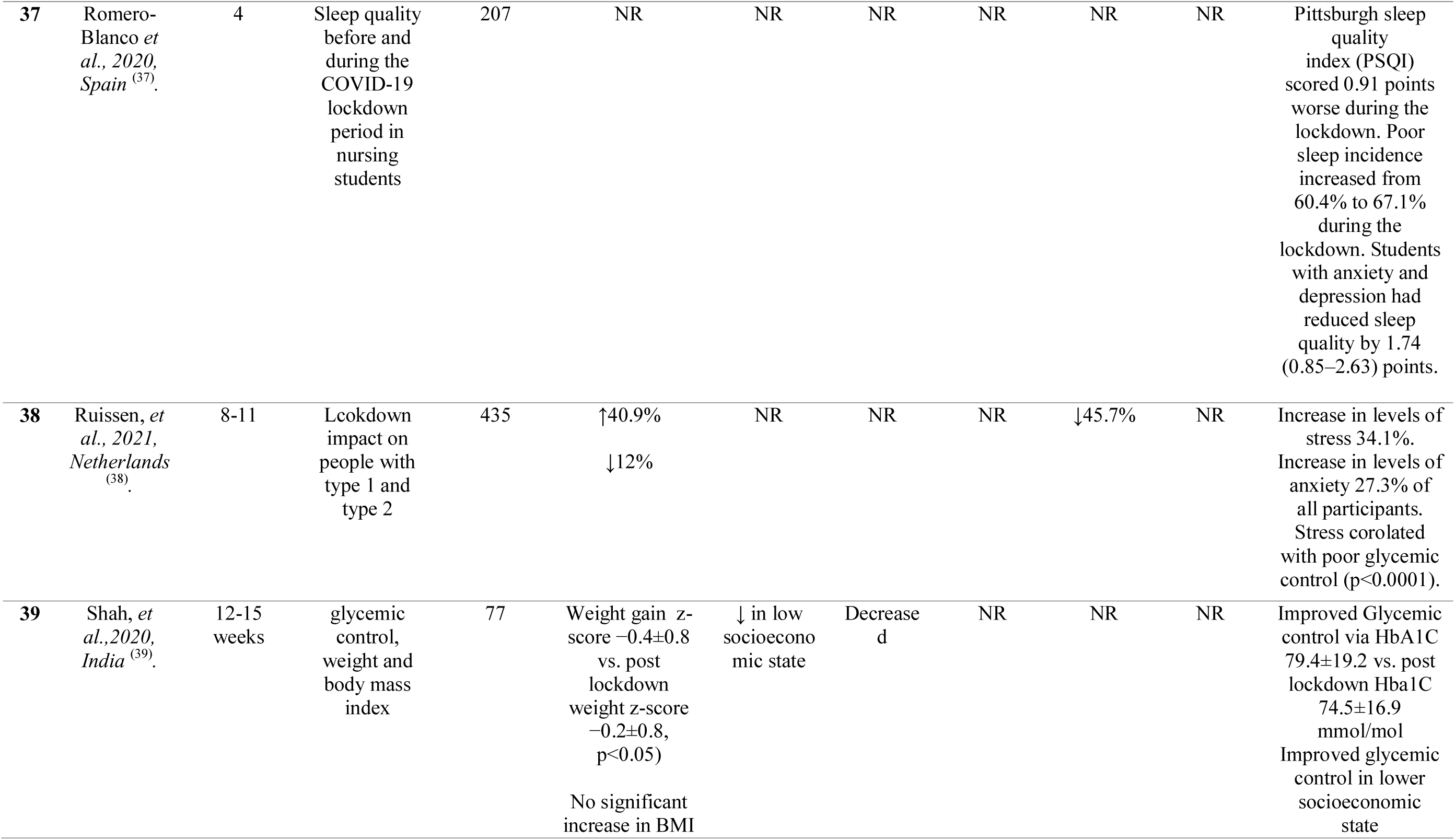
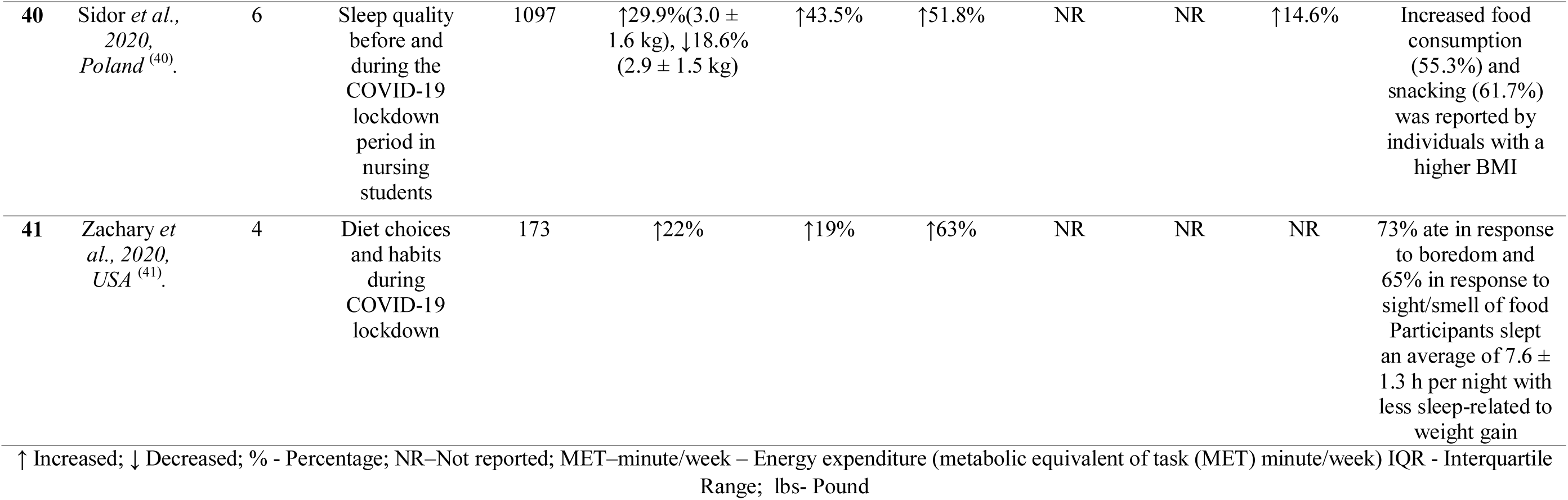
Behavioral and dietary changes related to pandemic confinements.

Appetite was modified either negatively or positively and was associated with employment change, suspension or working from home ^(21,52,53,58)^, or due to suspension of school attendance ^(24,48–50)^.

The initiating factors were as follows: response to smell and sight of food ^(25,26)^, boredom, binge eating and food cravings ^(25,26,41,43,45,55,67)^, snacking post dinner ^(26,33,34,39)^ and visual stimulation through social media^(33)^. A significant correlation was observed between snacking, the consumption of high density processed food, and a higher BMI ^(21,23,25,39)^. Increased energy intake by 10%–49.4% was observed among study participants ^(21,23,25,26,33–39,41,43–45,52,55,67)^, particularly those with an increased consumption of high density processed foods ^(21,23,24,35–37,39,41,43,44,51,68)^, female sex ^(21,35,39,43,52,64)^, or with a higher BMI ^(21,23–25,37,39,44,45,49,64)^. There was an increase in the number of meals eaten per day ^(24,36,37,45,68)^ and participants ate more than usual ^(35,39,43,55)^. The proportion of respondents engaged in cooking increased from 40% to 62% in our study sample ^(25,33,39,52)^. Likewise, consumption of homemade recipes increased ^(23,24,33,34,39,52)^ and eating homemade desserts increased compared to pre-lockdown^(21,23,24,33,35,39,68)^.

Less than one-third of the surveyed participants consumed fresh fruits and vegetables on a daily basis, while a similar number consumed sweets and desserts every day ^(21,22,25,33,37,39,40,52)^. In contrast, some studies have shown a decrease in unhealthy food consumption ^(23,34,39,55,68)^

Where Mediterranean diet was followed, 18- to 30-year-olds were more compliant than other age groups ^(21)^. Inverse associations were found between adherence to Mediterranean diet, and BMI ^(21,51)^. A total of 54% of respondents used leftovers for at least a third of meals, and those who shopped at farmers’ markets or local or organic markets ate up leftovers more (OR = 1.468, p < 0.001) ^(21)^. Among app users, mobile behavioral change app interaction was reduced by 9% ^(22)^. Eating in response to stress was associated with weight gain ^(26,35,59,67)^.

There was increased alcohol consumption ^(25,47,56,62,68)^ during the lockdown, while a decrease in alcohol consumption was also noted compared with pre-COVID-19 in another study ^(21,47)^. There was an increase in cigarette smoking generally ^(47,51,56,64)^ while in contrast, 3.3% of the smokers surveyed reported reduced smoking during quarantine ^(20)^.

Although, the participants reported spending more time in bed before lockdown ^(24,26,55,57)^, the overall sleep quality was worse ^(46,55,63)^. In contrast, secondary school students felt refreshed on awakening and increased sleeping hours ^(51)^. Weight gain was reported by others to be related to decreased night time sleep and reduced physical activity time ^(26,41,60)^

Sedentary lifestyle and screen time increased during the lockdown ^(24,26,38,47,55–57)^. Those participants who were not currently working or those who started working from home felt that they gained more weight compared with participants who did not have a change in job routine ^(21,21,52,58)^.

Physical activity altered by varying amounts, reduced in some studies to between 18% and 84% ^(23,24,34,35,40,47,52,54,55,59)^. People who were already overweight or obese engaged in less physical activity and had decreased energy expenditure during lockdown ^(37,39,44,52–54,56,59)^. Obese children spent less time participating in sports activities ^(24)^.

By contrast, studies reporting an increase in physical activity ^(21,52)^ found greater engagement in yoga/pilates, functional training, home training, and treadmill use and overall increased training frequency ^(21)^.

### Behavior changes observed in obese participants

Weight gain was more common in those already overweight or obese prior to lockdown and in individuals with pre-existing difficulty in weight management ^(21,23–25,37,39,44,46,49,64)^.

Increased snacking and food consumption were observed in participants with a higher BMI ^(24,25,33,34,68)^. Many of the participants agreed that they consumed less fruits and vegetables on a daily basis ^(22,25,34,52,69,70)^ but more high energy processed foods ^(23–25,41,44)^.

This intake was associated with an enhanced appetite and after-dinner hunger ^(21,37,39,45)^. Obese children reported an increase in the number of meals eaten along with an increased consumption of sweetened drinks, potato chips, and red meat ^(24)^. A decrease in intensive physical activity was associated with obesity ^(54)^. An inverse relationship was found between changes occurring in sporting activities and the number of meals consumed per day ^(24,41,53,68)^. The participants self-reporting anxiety and depression displayed an estimated weight gain ^(23,45,55,56,58,62)^.

### Determinants that can influence body weight during pandemics

Table 4 describes the determinants of body weight changes during the pandemic. Many determinants that can influence increased weight gain during confinement were identified via this current systematic review. This includes past behaviors, dietary behaviors, physical activity patterns, work environment, psychosocial and socioeconomic factors, and pre-existing comorbidities.

**Table 4:** Determinants of body weight during pandemic confinements.

| **Determinants that can have an effect on weight gain** |
| --- |
| **Demographics** |
| Female <sup>(1–6)</sup> |
| Baseline obese and overweight <sup>(2,3,6–15)</sup> |
| BMI <24 <sup>(16,17)</sup> |
| Age group > 45 years <sup>(3,9,18)</sup> |
| Age group < 25 years <sup>(1,9)</sup> |
| Having children under the age of 18 at home <sup>(1)</sup> |
| Changed work environment to working from home <sup>(1,2,19)</sup> |
| **Work Environment** |
| Loss of job <sup>(2)</sup> |
| Interruption of work routine <sup>(2)</sup> |
| Changed work habits: Furloughed or working from home<sup>(2)</sup> |
| Suspension of schools <sup>(17)</sup> |
| **Dietary Behaviors** |
| Increased food consumption <sup>(3,4,7,12,14,20–22)</sup> |
| Decreased consumption of fresh food products (particularly fruits, vegetables, and fish) <sup>(2,3,12,16,23)</sup> |
| Increased consumption of homemade recipes, sweets, and pizza <sup>(7,8)</sup> |
| Increased home cooking <sup>(3)</sup> |
| Increased cereal consumption <sup>(2,7)</sup> |
| Consumption of unhealthy foods <sup>(2–5,7,7,8,10,12,24,25)</sup> |
| Poor attention to diet balance <sup>(7)</sup> |
| Snacking after dinner <sup>(2,26)</sup> |
| Binge eating<sup>(24,27)</sup> |
| Loss of control to eating<sup>(24)</sup> |
| Eating in response to stress as a coping mechanism <sup>(23,26)</sup> |
| Eating secondary to appearance and smell of food <sup>(26)</sup> |
| Emotional eating <sup>(5,11,23)</sup> |
| Increase in alcohol intake <sup>(3,4,12)</sup> |
| **Psychological factors** |
| Decreased sleep time <sup>(22,26)</sup> |
| Lower sleep quality <sup>(28,29)</sup> |
| Stress <sup>(11,19,19,23,26,30–33)</sup> |
| Boredom <sup>(7)</sup> |
| Being alone <sup>(4,7)</sup> |
| Anxiety/Depression <sup>(6,7,19,24,32)</sup> |
| Depressive symptoms <sup>(1,4,11,24)</sup> |
| Mood disturbances <sup>(10)</sup> |
| Weight/ shape concerns <sup>(21)</sup> |
| **Socioeconomic factors** |
| Lack of garden <sup>(18)</sup> |
| Urban residence <sup>(1,32)</sup> |
| Lower education level <sup>(7,19)</sup> |
| Residence in a macroeconomic region >50% of EU-28 GDP<sup>(12)</sup> |
| Lower socioeconomic level <sup>(19,34)</sup> |
| **Physical inactivity** |
| Physical activity before lockdown <sup>(35)</sup> |
| Decreased physical activity <sup>(3–5,7,8,12,15,16,19–22,24,26,29–31,33,35–39)</sup> |
| Limitations of outdoor and in-gym activities <sup>(2,4,5,7)</sup> |
| Increased screen/tv time <sup>(7,12,21,31,33,36,39)</sup> |
| **Comorbidities** |
| Associated chronic illness <sup>(1,33,39–41)</sup> |
| **Determinants that can be associated with weight loss** |
| Underweight before confinement <sup>(9,12)</sup> |
| Younger age <sup>(12)</sup> |
| Remote work<sup>(12)</sup> |
| Urban residence <sup>(12,33)</sup> |
| Ate less <sup>(12)</sup> |
| Ate more fruits/vegetable <sup>(12)</sup> |
| Drank more water <sup>(12)</sup> |
| Ate more pulses/seafood/fish <sup>(12)</sup> |
| Did not consume alcohol <sup>(12)</sup> |
| Regular exercise before lockdown <sup>(6)</sup> |

Female sex ^(21,35,39,43,52,64)^, age under 25 years and over 45 years ^(25,39,52,54)^ are in particular at higher risk of gaining weight. Initial weight status, diet quality and physical exercise pattern before lockdown are important factors ^(21,23–25,37,45,49,52,64)^. In Chinese^(40)^ and Korean ^(50)^ populations, BMI <24 kg/m^2^ was associated with weight gain. However, some observed that those who were underweight before confinement lost more weight during confinement^(25,37)^. Poor diet quality before the lockdown was associated with weight gain ^(52)^. Decreased consumptions of legumes, fruits, and vegetables ^(25,39)^ was related to an increased consumption of sweets ^(23–25)^. Moreover, more home cooking with consumption of unhealthy foods are associated with increased weight gain^(20,22,23,34–36,38,40,42,43,48,65)^ as is increased alcohol intake ^(35,37,39,47,68)^.

Less intense physical behaviors were noted during lockdown periods compared with behaviors before lockdown casuing increased weight gain^(23,24,26,33–35,37–41,43,46,47,49,53,55–60)^.

This was due to the limitations of outdoor activities and in-gym activities^(21,43,53)^. In addition, there has been more sedentary behavior with increased screen time ^(23,37,38,47)^ which has been associated with weight gain.

Changed working habits; whether furloughed or working from home during the lockdown or those who had their job suspended ^(21,52,58)^, having children aged <18 years at home ^(52)^, urban residence and attaining a lower educational level ^(23)^ were associated with weight gain.

Patients with pre-existing psychiatric comorbidities had weight gain during COVID-19 lockdown ^(35,38,44,45,52,62)^, and stress ^(22,26,45,55,56,56,58,61,62)^, anxiety and/or depression^(23,58,61,62)^, eating in response to stress ^(22,26)^, boredom ^(23)^, living alone^(23)^ emotional eating (EE) ^(22,43,45)^ or weight or body shape concerns^(38)^ were associated with an increase in body weight during confinement. Decreased sleeping time ^(26)^ or poor quality sleep ^(46,51,63)^were further associated with weight gain.

Socio economic factors such as urban residence ^(52,61)^, lack of access to garden^(54)^, lower socio economic level ^(48)^ or lower education levels ^(23)^ and residence in a macroeconomic region ^(37)^ were associated with significant gain in the weight.

Patients with chronic illness such as diabetes, hypertension, lung disease, chronic coronary heart disease, congestive heart failure, depression or disability affecting one or more activities of daily living or lower levels of physically activity had an increase in weight ^(35,41,45,47,52,54,56,62,65)^.

Those who were previously underweight before the lockdown tended to lose more weight ^(25,25,37)^. Those whose diet included more fruits and vegetables, pulses and drank more water lost weight ^(37)^.

## Discussion

This systematic review highlights contrasting effects of pandemic confinements on body weight, and we identified specific factors associated with change in body weight during the lockdown periods.

A BMI of >25 kg/m^2^ was identified as an independent risk factor for increased food intake during lockdown ^(71)^. Other influences were inadequate sleep, decreased physical activity, EE in response to stress, lack of control in dietary habits ^(21,25,54)^, and increased alcohol consumption and smoking ^(35,37,39,51,56,64,68)^. The impact of these influences is more significant in the obese population.

Eating habits as well as diet composition are linked to weight gain ^(72)^. Increased snacking after meals, particularly post dinner, was associated with weight gain ^(72)^. Jakubowicz et al. also concluded that increased calories at dinner increased the subjects’ weight ^(73)^. Thus, decreasing food consumption during and post dinner should be recommended.

Social networks, neighborhood social activities, and physical activity can influence an individual’s opportunity to make better choices contributing to protection from obesity ^(74)^. The absence of these influences during extended lockdown periods may facilitates a more obesogenic environment, thus encouraging weight gain ^(75)^.

By contrast, not all effects of pandemic confinement resulted in weight gain. In an Italian study, 38% of participants adhered to a Mediterranean diet. This may have been assisted by the Italian Ministry of Health publishing online materials regarding favorable lifestyle choices during the lockdown in April 2020 and providing practical guidelines on healthy behaviors ^(76,77)^.

Pandemic confinements undoubtedly increase stress^(34,34,56,59–61,61)^, 73% and 83% of respondents experienced an increase in anxiety and depression, respectively, with 70% reporting weight management issues, stock-piling food, and stress eating ^(23,41,78)^. Weight loss was reported in three studies by 13% to 19% of participants ^(21,25,52)^. Two studies showed stress related weight among working professionals and university students ^(79,80)^. The mechanism is twofold and results from decreased, unchanged or increased energy intake coupled with adaptive adrenergic stimulated thermogenesis involving brown adipose tissues ^(81)^. The weight loss observed in this systematic review may also be attributed to the negative effect of stress ^(21,25,26,52,82)^.

The link between weight changes and stress has been studied extensively ^(83,84)^. Behavioral and physiological explanations suggest that the sensation of eating is associated with a psychological escape from emotional distress ^(85)^ and that the consumption of high calorie foods alleviates stress ^(83)^. During a pandemic, where cities and even entire nations were locked down, fear and anxiety related to COVID-19 induced an over eating behavior. However, management of this associated condition is difficult ^(86)^. The adverse effects of lockdown on the psychological and social wellbeing of society emphasize the need for strong public health interventions to support particularly at-risk people.

The associations between health outcomes, exercise, and physical activity are well-established. The results from studies that we included in this review were mixed; some participants engaged in increased physical activity, while others had lower levels of physical activity. Confinement did not induce many sedentary participants to increase their physical activity. Other unhealthy behaviors such as increased screen time were noted which are similar to previous studies^(87)^. Stress may impair efforts to become physically active; conversely, those who already participate may do so to reduce stress ^(88)^, which may explain the variation in physical activity observed. Siegel et al. (2002) describes this as stress-related behavioral activation or inhibition ^(89)^.

Other unhealthy behaviors were noted during the confinement. There was a 14.6% increase in the consumption of alcohol in participants who had issues with alcohol ^(25)^. In the acute post-disaster period of the September 11 attacks in Manhattan, New York City, the prevalence of alcohol consumption and marijuana use among New York City residents increased over a five- to eight-week period ^(90)^. These results mirror our findings, suggesting shared responses to intense community stresses. Although these activities may not directly affect weight, alcohol consumption and obesity are common risk factors for chronic illnesses leading to increased morbidity and mortality ^(91)^. Furthermore, in a study conducted in the Netherlands, it was reported that overweight and obese individuals found it more difficult to make healthy food choices. More savory snacks and non-alcoholic beverages were purchased and consumed at home (35.6%) because of more leisure time (31.5%) and boredom (21.9%) during the lockdown ^(92)^.

Positive outcomes from confinement have also been reported ^(93)^. These behaviors may result from the increased availability of time to cook, health risk perceptions, lack of negative social distractions ^(94)^, and socio-cognitive ideation toward a healthier lifestyle ^(95)^. Long-term studies are necessary to determine whether these constructive and preventive behaviors can be sustained after confinement is over.

Food security, which involves food availability, accessibility, and affordability, is another important factor in the relationship between pandemic confinement and body weight changes ^(96)^. Global non-pharmaceutical interventions, such as lockdowns and quarantines, implemented to limit the spread of the virus have seriously impacted food security systems ^(97)^, with the greatest burden affecting communities in which nutritional health is fragile ^(98)^. Communities with precarious budgeting practices were destabilized by food price inflation and product shortages. Additional influences on food security included movement restrictions of workers, changes in consumer demand, closure of food production facilities, restricted food trade policies, and financial pressures in the food supply chain. As dependence on food banks grew with an exponential increase in demand, basic survival needs presided over healthy dietary choices ^(18)^. Prior to 2020, 690 million people were already food-insecure and hungry^(99)^. By the end of 2020, the COVID-19 pandemic had created an additional 270 million food-insecure people ^(100,101)^. Unfortunately, vulnerable populations are not restricted to under-resourced countries; developed nations are suffering as well. In the United States alone, food insecurity more than doubled as a result of the economic crisis brought on by the outbreak, impacting as many as 23% of households ^(102)^.

Serious ethical and health-related issues hinder healthcare providers working with vulnerable populations. In general, differences in weight status and dietary intake reveal that a trend in obesity increases as the degree of food insecurity increases ^(103)^. The COVID-19 crisis has highlighted food insecurity as a significant factor in nutritional poverty ^(97)^. This awareness of food insecurity may provide nations with the impetus to robustly tackle food-related epidemics, such as obesity and diabetes.

COVID-19 has challenged us to consider the role and balance of healthcare, personal health, and holistic wellbeing. Redefining these dynamics in preparation for future pandemics is imperative to minimize severe impacts to health and resources ^(104)^. It was previously observed that consumerism is affected by internal factors, such as personal character, and external factors, such as economic crises. The pandemic served as an external factor that altered consumer behavior ^(105)^.

Relief efforts by governmental and non-governmental agencies achieved temporary solutions without significant public pressure ^(106)^, but the demand for aid from all sectors of society is mounting. National governments should take the lead in providing strategic directions that will ensure the continuity of food accessibility to all, particularly the most vulnerable. Focus must be on coordinated and integrated public health programs through legislative action to end sub-standard dietary conditions endured by those most in need. By collaborating with key stakeholders, health professionals must provide aggressive nutritional counseling to improve dietary habits, and concerted efforts across the board are paramount.

Recent research has shown obesity to be an independent risk factor for severe complications and increased mortality from COVID-19 ^(107,108)^. The evidence suggests a linear relationship with obesity increasing the risk of severe disease and death among COVID-19 patients ^(109)^. The co-existence of both pandemics, COVID-19 and obesity, along with the emergence of obesity evolving from lockdown have caused a ‘syndemic’ or a symbiotic pandemic ^(110)^. Researchers must address the significant knowledge gaps that have become apparent during this pandemic regarding preparedness and response to such a crisis. Moreover, COVID-19 has disproportionately affected certain populations, and future research should focus on such vulnerable populations to ensure better outcomes.

### Strengths and limitations

To our knowledge, this is the first systematic review evaluating the effects of pandemic confinement on body weight. Our study highlights major determinants that can have an impact on body weight during confinement and those that can be targeted in future pandemics to effectively manage body weight during pandemics via public health initiatives. Moreover, confinements are not solely related to pandemics and can also occur during natural disasters or calamities and in prisons. Determinants identified could be modified via appropriate public health measures to reduce negative impacts.

This study has limitations. First, there was limited evidence from past pandemics related to obesity and morbidity or mortality. This may reflect the recent evolution of worldwide obesity ^(111)^. Second, within the common research theme of body weight changes during pandemic confinements, our systematic review found marked heterogeneity in the determinants and measured outcomes. This variation could be explained by differences in the study population and types of outcome measurements ^(112)^. Nevertheless, in our systematic review, we followed a rigorous protocol with clear objectives and inclusion and exclusion criteria. This allowed for the identification and pooling of the determinants of body weight changes during pandemic confinements (Table 4). A thorough and complete identification of the different determinants related to pandemic confinements could guide decision makers. Furthermore, our study calls for further research into the level of impact of each determinant. Third, given the contemporary nature of the pandemic, the literature was primarily related to countries where COVID-19 had an early “first wave” impact. Findings from other continents, particularly from Africa and South America, are yet to emerge. Fourth, online surveys using social media platforms were the predominant data collection method, which has recognized strengths and biases. Although the researchers used this form of data collection to reach a wider population, the likelihood of a bias toward a younger population should be noted. Fifth, although this analysis provides evidence for the effects of confinement on body weight, we are unable to comment on the potential for interventions such as lifestyle changes to attenuate the phenomenon. Sixth, because of the limited number of studies included, we were unable to correct for influences, such as preexisting diets, and could not quantify the impact of possible factors in isolation. Although we know that weight gain is likely during confinement, further research using more sophisticated data collection techniques is necessary to determine the holistic impact of confinement to provide evidence-based practical solutions for future eventualities.

## Conclusion

This systematic review highlights the significant effects that pandemic confinements can have in the short term on body mass. Poor sleep, snacking post dinner, lack of dietary restraint, pre-existing overweight status, EE due to stress, and decreased physical activity are risk factors for weight gain.

Preparing for the next “wave” is challenging given the multitude of factors that must be tailored to the local situations and available resources. Planning for future episodes requires a strong, evidence-based national policy in conjunction with clear guidelines to ensure that the negative sequelae of lockdowns are minimized.

## Data Availability

Not applicable

## Ethical statement

This article was not plagiarized and had not previously been published in other journals.

## Acknowledgements

We thank Gamila Hassan at the National Medical Library at UAEU for her strategic support in locating and uploading full-text articles to Covidence.

## Authorship

MK, PM, RG, LÖ, and HM formulated the research question and designed the study MK, PM, RG, KA and AS extracted and reviewed the data independently. LÖ and MK performed the literature search. MK, PM, RG, AS, and JN performed the literature review and data analysis. MK, PM, KA, HM, RG, AS, JN, LÖ, JS, EMS and JK contributed to drafting the paper. MK, RG, PM, AS, KA equally contributed to all of the work as co-first authors.

## References

1. CDC COVID-19 Response Team (2020) Severe Outcomes Among Patients with Coronavirus Disease 2019 (COVID-19) - United States, February 12-March 16, 2020. MMWR Morb. Mortal. Wkly. Rep. 69, 343–346.

2. Madhav N, Oppenheim B, Gallivan M, et al. (2017) Pandemics: Risks, Impacts, and Mitigation. In Disease Control Priorities: Improving Health and Reducing Poverty, 3rd ed. [Jamison DT, Gelband H, Horton S, et al., editors]. Washington (DC): The International Bank for Reconstruction and Development / The World Bank.

3. Ismail L, Materwala H, Znati T, et al. (2020) Tailoring time series models for forecasting coronavirus spread: Case studies of 187 countries. Computational and Structural Biotechnology Journal 18, 2972–3206.

4. Khan MA & Moverley Smith JE (2020) “Covibesity,” a new pandemic. Obesity Medicine 19, 100282.

5. Al Falasi RJ & Ab Khan M (2020) The impact of COVID-19 on Abu Dhabi and its primary care response. Australian journal of general practice 49.

6. Every-Palmer S, Jenkins M, Gendall P, et al. (2020) Psychological distress, anxiety, family violence, suicidality, and wellbeing in New Zealand during the COVID-19 lockdown: A cross-sectional study. PLoS one 15, e0241658. Public Library of Science San Francisco, CA USA.

7. Mozaffarian D, Hao T, Rimm EB, et al. (2011) Changes in Diet and Lifestyle and Long-Term Weight Gain in Women and Men. N Engl J Med 364, 2392–2404.

8. Khan MA, Hashim MJ, Mustafa H, et al. (2020) Global Epidemiology of Ischemic Heart Disease: Results from the Global Burden of Disease Study. Cureus 12, e9349.

9. Gallo L, Moritz K & Akison L (2020) Nutrient Intake, Physical Activity Levels, and Metabolic Status in Australian University Biomedical Students. Curr Dev Nutr 4, 1404–1404. Oxford Academic.

10. Tsenoli M, Moverley Smith JE & Khan MA (2021) A community perspective of COVID-19 and obesity in children: Causes and consequences. Obesity Medicine 22, 100327.

11. Diaz-Zavala RG, Castro-Cantú MF, Valencia ME, et al. (2017) Effect of the Holiday Season on Weight Gain: A Narrative Review. J Obes 2017.

12. Díaz-Zavala RG, Castro-Cantú MF, Valencia ME, et al. (2017) Effect of the Holiday Season on Weight Gain: A Narrative Review. J Obes 2017.

13. Rundle AG, Park Y, Herbstman JB, et al. (2020) COVID-19-Related School Closings and Risk of Weight Gain Among Children. Obesity (Silver Spring) 28, 1008–1009.

14. Mason F, Farley A, Pallan M, et al. (2018) Effectiveness of a brief behavioural intervention to prevent weight gain over the Christmas holiday period: randomised controlled trial. BMJ 363, k4867.

15. Hafner JW, Hough SM, Getz MA, et al. (2010) All-terrain vehicle safety and use patterns in Central Illinois youth. The Journal of Rural Health 26, 67–72. Wiley-Blackwell Publishing Ltd.

16. Pearl RL (2020) Weight Stigma and the ‘Quarantine-15’. Obesity (Silver Spring) 28, 1180–1181.

17. Laborde D, Martin W, Swinnen J, et al. (2020) COVID-19 risks to global food security. Science 369, 500–502. American Association for the Advancement of Science.

18. Huizar MI, Arena R & Laddu DR (2020) The global food syndemic: The impact of food insecurity, Malnutrition and obesity on the healthspan amid the COVID-19 pandemic. Prog Cardiovasc Dis, S0033-0620(20)30139–0. Elsevier Inc.

19. The Lancet Global Health (2020) Food insecurity will be the sting in the tail of COVID-19. Lancet Glob Health 8, e737–e737. The Author(s). Published by Elsevier Ltd.

20. Cuschieri S & Grech S (2020) Obesity population at risk of COVID-19 complications. Global health, epidemiology and genomics 5. Cambridge University Press.

21. Di Renzo L, Gualtieri P, Pivari F, et al. (2020) Eating habits and lifestyle changes during COVID-19 lockdown: an Italian survey. Journal of Translational Medicine 18, 1–15. BioMed Central.

22. Mitchell ES, Yang Q, Behr H, et al. (2020) Self-reported food choices before and during COVID-19 lockdown. medRxiv, 2020.06.15.20131888. Cold Spring Harbor Laboratory Press.

23. Pellegrini M, Ponzo V, Rosato R, et al. (2020) Changes in Weight and Nutritional Habits in Adults with Obesity during the ‘Lockdown’ Period Caused by the COVID-19 Virus Emergency. Nutrients 12.

24. Pietrobelli A, Pecoraro L, Ferruzzi A, et al. (2020) Effects of COVID-19 lockdown on lifestyle behaviors in children with obesity living in Verona, Italy: a longitudinal study. Obesity. Wiley Online Library.

25. Sidor A & Rzymski P (2020) Dietary Choices and Habits during COVID-19 Lockdown: Experience from Poland. Nutrients 12, 1657. Multidisciplinary Digital Publishing Institute.

26. Zachary Z, Brianna F, Brianna L, et al. (2020) Self-quarantine and Weight Gain Related Risk Factors During the COVID-19 Pandemic. Obesity Research & Clinical Practice. Elsevier.

27. Higgins JP, Thomas J, Chandler J, et al. (2019) Cochrane handbook for systematic reviews of interventions. John Wiley & Sons.

28. Moher D, Liberati A, Tetzlaff J, et al. (2009) Preferred reporting items for systematic reviews and meta-analyses: the PRISMA statement. Ann. Intern. Med. 151, 264–269, W64.

29. Zhang J, Zhang Y, Huo S, et al. (2020) Emotional Eating in Pregnant Women during the COVID-19 Pandemic and Its Association with Dietary Intake and Gestational Weight Gain. Nutrients 12.

30. Herzog R, Álvarez-Pasquin MJ, Díaz C, et al. (2013) Are healthcare workers’ intentions to vaccinate related to their knowledge, beliefs and attitudes? A systematic review. BMC public health 13, 154. Springer.

31. Modesti PA, Reboldi G, Cappuccio FP, et al. (2016) Panethnic Differences in Blood Pressure in Europe: A Systematic Review and Meta-Analysis. PLOS ONE 11, e0147601. Public Library of Science.

32. Peterson J, Welch V, Losos M, et al. (2011) The Newcastle-Ottawa scale (NOS) for assessing the quality of nonrandomised studies in meta-analyses. Ottawa: Ottawa Hospital Research Institute.

33. Cheikh Ismail L, Osaili TM, Mohamad MN, et al. (2020) Eating habits and lifestyle during COVID-19 lockdown in the United Arab Emirates: a cross-sectional study. Nutrients 12, 3314. Multidisciplinary Digital Publishing Institute.

34. Ismail LC, Osaili TM, Mohamad MN, et al. (2020) Assessment of Eating Habits and Lifestyle during Coronavirus Pandemic in the MENA region: A Cross-Sectional Study. British Journal of Nutrition, 1–30. Cambridge University Press.

35. Di Santo SG, Franchini F, Filiputti B, et al. (2020) The effects of COVID-19 and quarantine measures on the lifestyles and mental health of people over 60 at increased risk of dementia. Frontiers in Psychiatry 11. Frontiers Media SA.

36. Dondi A, Candela E, Morigi F, et al. (2021) Parents’ Perception of Food Insecurity and of Its Effects on Their Children in Italy Six Months after the COVID-19 Pandemic Outbreak. Nutrients 13, 121. Multidisciplinary Digital Publishing Institute.

37. Drywień ME, Hamulka J, Zielinska-Pukos MA, et al. (2020) The COVID-19 Pandemic Lockdowns and Changes in Body Weight among Polish Women. A Cross-Sectional Online Survey PLifeCOVID-19 Study. Sustainability 12, 7768. Multidisciplinary Digital Publishing Institute.

38. Keel PK, Gomez MM, Harris L, et al. (2020) Gaining “The Quarantine 15:” Perceived versus observed weight changes in college students in the wake of COVID-19. International Journal of Eating Disorders 53, 1801–1808. Wiley Online Library.

39. Kriaucioniene V, Bagdonaviciene L, Rodríguez-Pérez C, et al. (2020) Associations between changes in health behaviours and body weight during the COVID-19 quarantine in Lithuania: the Lithuanian COVIDiet Study. Nutrients 12, 3119. Multidisciplinary Digital Publishing Institute.

40. He M, Xian Y, Lv X, et al. (2020) Changes in Body Weight, Physical Activity, and Lifestyle During the Semi-lockdown Period After the Outbreak of COVID-19 in China: An Online Survey. Disaster Med Public Health Prep, 1–6.

41. Athanasiadis DI, Hernandez E, Hilgendorf W, et al. (2020) How are bariatric patients coping during the coronavirus disease 2019 (COVID-19) pandemic? Analysis of factors known to cause weight regain among postoperative bariatric patients. Surgery for Obesity and Related Diseases. Elsevier.

42. Błaszczyk-Bębenek E, Jagielski P, Bolesławska I, et al. (2020) Nutrition Behaviors in Polish Adults before and during COVID-19 Lockdown. Nutrients 12, 3084. Multidisciplinary Digital Publishing Institute.

43. Ozden G & Parlar Kiliç S (2021) The Effect of Social Isolation during COVID-19 Pandemic on Nutrition and Exercise Behaviors of Nursing Students. Ecology of Food and Nutrition, 1–19. Taylor & Francis.

44. Jimenez A, de Hollanda A, Palou E, et al. (2021) Psychosocial, Lifestyle, and Body Weight Impact of COVID-19-Related Lockdown in a Sample of Participants with Current or Past History of Obesity in Spain. Obesity surgery, 1–10. Springer.

45. Marchitelli S, Mazza C, Lenzi A, et al. (2020) Weight Gain in a Sample of Patients Affected by Overweight/Obesity with and without a Psychiatric Diagnosis during the Covid-19 Lockdown. Nutrients 12, 3525. Multidisciplinary Digital Publishing Institute.

46. Martínez-de-Quel Ó, Suárez-Iglesias D, López-Flores M, et al. (2021) Physical activity, dietary habits and sleep quality before and during COVID-19 lockdown: A longitudinal study. Appetite 158, 105019. Elsevier.

47. Chagué F, Boulin M, Eicher J-C, et al. (2020) Impact of lockdown on patients with congestive heart failure during the coronavirus disease 2019 pandemic. ESC Heart Fail.

48. Shah N, Karguppikar M, Bhor S, et al. (2020) Impact of lockdown for COVID-19 pandemic in Indian children and youth with type 1 diabetes from different socio-economic classes. Journal of Pediatric Endocrinology and Metabolism 1. De Gruyter.

49. Adıbelli D & Sümen A (2020) The effect of the coronavirus (COVID-19) pandemic on health-related quality of life in children. Children and Youth Services Review 119, 105595.

50. Kang HM, Jeong DC, Suh B-K, et al. (2021) The Impact of the Coronavirus Disease-2019 Pandemic on Childhood Obesity and Vitamin D Status. J Korean Med Sci 36. The Korean Academy of Medical Sciences.

51. Dragun R, Veček NN, Marendić M, et al. (2021) Have Lifestyle Habits and Psychological Well-Being Changed among Adolescents and Medical Students Due to COVID-19 Lockdown in Croatia? Nutrients 13, 97. Multidisciplinary Digital Publishing Institute.

52. Deschasaux-Tanguy M, Druesne-Pecollo N, Esseddik Y, et al. (2020) Diet and physical activity during the COVID-19 lockdown period (March-May 2020): results from the French NutriNet-Sante cohort study. medRxiv, 2020.06.04.20121855. Cold Spring Harbor Laboratory Press.

53. Giustino V, Parroco AM, Gennaro A, et al. (2020) Physical Activity Levels and Related Energy Expenditure during COVID-19 Quarantine among the Sicilian Active Population: A Cross-Sectional Online Survey Study. Sustainability 12, 4356. Multidisciplinary Digital Publishing Institute.

54. Rogers N, Roberts C, Waterlow N, et al. (2020) Behavioural change towards reduced intensity physical activity is disproportionately prevalent among adults with serious health issues or self-perception of high risk during the UK COVID-19 lockdown. London School of Hygiene & Tropical Medicine.

55. Chopra S, Ranjan P, Singh V, et al. (2020) Impact of COVID-19 on lifestyle-related behaviours-a cross-sectional audit of responses from nine hundred and ninety-five participants from India. Diabetes & Metabolic Syndrome: Clinical Research & Reviews 14, 2021–2030. Elsevier.

56. Cransac-Miet A, Zeller M, Chagué F, et al. (2021) Impact of COVID-19 lockdown on lifestyle adherence in stay-at-home patients with chronic coronary syndromes: Towards a time bomb. International Journal of Cardiology 323, 285–287. Elsevier.

57. Jia P, Zhang L, Yu W, et al. (2020) Impact of COVID-19 lockdown on activity patterns and weight status among youths in China: the COVID-19 Impact on Lifestyle Change Survey (COINLICS). International Journal of Obesity, 1–5. Nature Publishing Group.

58. Malkawi SH, Almhdawi K, Jaber AF, et al. (2020) COVID-19 Quarantine-Related Mental Health Symptoms and their Correlates among Mothers: A Cross Sectional Study. Maternal and Child Health Journal.

59. de Matos DG, Aidar FJ, Almeida-Neto PF de, et al. (2020) The impact of measures recommended by the government to limit the spread of coronavirus (COVID-19) on physical activity levels, quality of life, and mental health of Brazilians. Sustainability 12, 9072. Multidisciplinary Digital Publishing Institute.

60. Ruissen MM, Regeer H, Landstra CP, et al. (2021) Increased stress, weight gain and less exercise in relation to glycemic control in people with type 1 and type 2 diabetes during the COVID-19 pandemic. BMJ Open Diab Res Care 9, e002035.

61. Ahmed HO (2020) The impact of social distancing and self-isolation in the last corona COVID-19 outbreak on the body weight in Sulaimani governorate-Kurdistan/Iraq, a prospective case series study. Annals of Medicine and Surgery 59, 110–117. Elsevier.

62. Gentile A, Torales J, O’Higgins M, et al. (2020) Phone-based outpatients’ follow-up in mental health centers during the COVID-19 quarantine. International Journal of Social Psychiatry, 0020764020979732. SAGE Publications Sage UK: London, England.

63. Romero-Blanco C, Rodríguez-Almagro J, Onieva-Zafra MD, et al. (2020) Sleep Pattern Changes in Nursing Students during the COVID-19 Lockdown. Int J Environ Res Public Health 17.

64. Dogas Z, Kalcina LL, Dodig IP, et al. (2020) The effect of COVID-19 lockdown on lifestyle and mood in Croatian general population: a cross-sectional study. Croatian medical journal 61, 309. Medicinska Naklada.

65. Onmez A, Gamsızkan Z, Özdemir Ş, et al. (2020) The effect of COVID-19 lockdown on glycemic control in patients with type 2 diabetes mellitus in Turkey. Diabetes & Metabolic Syndrome: Clinical Research & Reviews 14, 1963–1966. Elsevier.

66. Karatas S, Yesim T & Beysel S (2021) Impact of lockdown COVID-19 on metabolic control in type 2 diabetes mellitus and healthy people. Primary Care Diabetes. Elsevier.

67. Mason TB, Barrington-Trimis J & Leventhal AM (2021) Eating to Cope With the COVID-19 Pandemic and Body Weight Change in Young Adults. Journal of Adolescent Health 68, 277–283. Elsevier.

68. Blaszczyk-Bębenek E, Jagielski P, Bolesławska I, et al. (2020) Nutrition behaviors in Polish adults before and during covid-19 lockdown. Nutrients 12, 3084. Multidisciplinary Digital Publishing Institute.

69. Athanasiadis DI, Hernandez E, Hilgendorf W, et al. (2020) How are bariatric patients coping during the coronavirus disease 2019 (COVID-19) pandemic? Analysis of factors known to cause weight regain among postoperative bariatric patients. Surgery for Obesity and Related Diseases. Elsevier.

70. Kriaucioniene V, Bagdonaviciene L, Rodríguez-Pérez C, et al. (2020) Associations between changes in health behaviours and body weight during the COVID-19 quarantine in Lithuania: the Lithuanian COVIDiet Study. Nutrients 12, 3119. Multidisciplinary Digital Publishing Institute.

71. Huber BC, Steffen J, Schlichtiger J, et al. (2020) Altered nutrition behavior during COVID-19 pandemic lockdown in young adults. European journal of nutrition, 1–10. Springer.

72. Newman E, O’Connor DB & Conner M (2007) Daily hassles and eating behaviour: the role of cortisol reactivity status. Psychoneuroendocrinology 32, 125–132.

73. Jakubowicz D, Barnea M, Wainstein J, et al. (2013) High caloric intake at breakfast vs. dinner differentially influences weight loss of overweight and obese women. Obesity (Silver Spring) 21, 2504–2512.

74. McNeill LH, Kreuter MW & Subramanian SV (2006) Social environment and physical activity: a review of concepts and evidence. Soc Sci Med 63, 1011–1022.

75. Swinburn B, Egger G & Raza F (1999) Dissecting obesogenic environments: the development and application of a framework for identifying and prioritizing environmental interventions for obesity. Prev Med 29, 563–570.

76. Gallè F, Sabella EA, Da Molin G, et al. (2020) Understanding Knowledge and Behaviors Related to CoViD-19 Epidemic in Italian Undergraduate Students: The EPICO Study. Int J Environ Res Public Health 17.

77. Salute M della Covid-19, how to follow an appropriate and healthy lifestyle when staying at home. http://www.salute.gov.it/portale/nuovocoronavirus/dettaglioNotizieNuovoCoronavirus. jsp?lingua=italiano&menu=notizie&p=dalministero&id=4421 (accessed July 2020).

78. Almandoz JP, Xie L, Schellinger JN, et al. (2020) Impact of COVID-19 stay-at-home orders on weight-related behaviours among patients with obesity. *Clin Obes*, e12386.

79. Kivimäki M, Head J, Ferrie JE, et al. (2006) Work stress, weight gain and weight loss: evidence for bidirectional effects of job strain on body mass index in the Whitehall II study. Int J Obes (Lond) 30, 982–987.

80. Serlachius A, Hamer M & Wardle J (2007) Stress and weight change in university students in the United Kingdom. Physiol. Behav. 92, 548–553.

81. Razzoli M & Bartolomucci A (2016) The Dichotomous Effect of Chronic Stress on Obesity. Trends Endocrinol. Metab. 27, 504–515.

82. Dallman MF (2010) Stress-induced obesity and the emotional nervous system. Trends in Endocrinology & Metabolism 21, 159–165. Elsevier.

83. Adam TC & Epel ES (2007) Stress, eating and the reward system. Physiol. Behav. 91, 449–458.

84. O’Connor DB, Jones F, Conner M, et al. (2008) Effects of daily hassles and eating style on eating behavior. Health Psychol 27, S20–31.

85. Heatherton TF & Baumeister RF (1991) Binge eating as escape from self-awareness. Psychol Bull 110, 86–108.

86. Haddad C, Kheir MB, Zakhour M, et al. (2020) Association between eating behavior and quarantine/confinement stressors during the Coronavirus disease 2019 outbreak..

87. Khan MA, Shah SM, Shehab A, et al. (2019) Screen time and metabolic syndrome among expatriate adolescents in the United Arab Emirates. Diabetes & Metabolic Syndrome: Clinical Research & Reviews 13, 2565–2569. Elsevier.

88. Stults-Kolehmainen MA & Sinha R (2014) The effects of stress on physical activity and exercise. Sports Med 44, 81–121.

89. Seigel K, Broman J-E & Hetta J (2002) Behavioral activation or inhibition during emotional stress-implications for exercise habits and emotional problems among young females. Nord J Psychiatry 56, 441–446.

90. Vlahov D, Galea S, Resnick H, et al. (2002) Increased use of cigarettes, alcohol, and marijuana among Manhattan, New York, residents after the September 11th terrorist attacks. Am. J. Epidemiol. 155, 988–996.

91. Chiolero A, Faeh D, Paccaud F, et al. (2008) Consequences of smoking for body weight, body fat distribution, and insulin resistance. Am. J. Clin. Nutr. 87, 801–809.

92. Poelman MP, Gillebaart M, Schlinkert C, et al. (2020) Eating behavior and food purchases during the COVID-19 lockdown: A cross-sectional study among adults in the Netherlands. Appetite 157, 105002. Elsevier.

93. Ruiz-Roso MB, de Carvalho Padilha P, Mantilla-Escalante DC, et al. (2020) Covid-19 Confinement and Changes of Adolescent’s Dietary Trends in Italy, Spain, Chile, Colombia and Brazil. Nutrients 12.

94. Ferrer R & Klein WM (2015) Risk perceptions and health behavior. Curr Opin Psychol 5, 85–89.

95. Raude J (2020) Determinants of preventive behaviors in response to the COVID-19 pandemic in France: comparing the sociocultural, psychosocial and social cognitive explanations. PsyArXiv.

96. Adams EL, Caccavale LJ, Smith D, et al. (2020) Food insecurity, the home food environment, and parent feeding practices in the era of COVID-19. Obesity 28, 2056–2063. Wiley Online Library.

97. Micha R, Mannar V, Afshin A, et al. (2020) 2020 Global nutrition report: action on equity to end malnutrition. Development Initiatives.

98. UNSCN (2020) COVID-19 pandemic: The evolving impact on how people meet the food system. https://www.unscn.org/en/news-events/recent-news?idnews=2065 (accessed January 2021).

99. As more go hungry and malnutrition persists, achieving Zero Hunger by 2030 in doubt, UN report warns. https://www.who.int/news/item/13-07-2020-as-more-go-hungry-and-malnutrition-persists-achieving-zero-hunger-by-2030-in-doubt-un-report-warns (accessed February 2021).

100. (2020) WFP at a glance. United Nations World Food Programme. https://www.wfp.org/stories/wfp-glance (accessed December 2020).

101. Paslakis G, Dimitropoulos G & Katzman DK (2021) A call to action to address COVID-19–induced global food insecurity to prevent hunger, malnutrition, and eating pathology. Nutrition Reviews 79, 114–116.

102. Schanzenbach D & Pitts A (2020) How much has food insecurity risen? Evidence from the Census Household Pulse Survey. Institute for Policy Research (IPR) Rapid Research Report. Northwestern Institute for Policy Research. Published June 10.

103. El Zein A, Colby SE, Zhou W, et al. (2020) Food insecurity is associated with increased risk of obesity in US college students. Current Developments in Nutrition 4, nzaa120. Oxford University Press.

104. Sheth J (2020) Impact of Covid-19 on Consumer Behavior: Will the Old Habits Return or Die? Journal of Business Research. Elsevier.

105. Mehta S, Saxena T & Purohit N (2020) The New Consumer Behaviour Paradigm amid COVID-19: Permanent or Transient? Journal of Health Management 22, 291–301. SAGE Publications Sage India: New Delhi, India.

106. Baraniuk C (2020) Fears grow of nutritional crisis in lockdown UK. bmj 370. British Medical Journal Publishing Group.

107. Hajifathalian K, Kumar S, Newberry C, et al. (2020) Obesity is associated with worse outcomes in COVID-19: Analysis of Early Data From New York City. Obesity (Silver Spring).

108. Ho FK, Celis-Morales CA, Gray SR, et al. (2020) Modifiable and non-modifiable risk factors for COVID-19: results from UK Biobank. medRxiv, 2020.04.28.20083295. Cold Spring Harbor Laboratory Press.

109. Huang Y, Yao LU, Huang Y-M, et al. (2020) Obesity in patients with COVID-19: a systematic review and meta-analysis. Metabolism, 154378. Elsevier.

110. Hill MA, Sowers JR & Mantzoros CS (2020) Commentary: COVID-19 and obesity pandemics converge into a syndemic requiring urgent and multidisciplinary action. Metabolism 114, 154408. Elsevier.

111. Eknoyan G (2006) A history of obesity, or how what was good became ugly and then bad. Adv Chronic Kidney Dis 13, 421–427.

112. Gagnier JJ, Moher D, Boon H, et al. (2012) Investigating clinical heterogeneity in systematic reviews: a methodologic review of guidance in the literature. BMC medical research methodology 12, 111. Springer.

## References

1. Adıbelli D & Sümen A (2020) The effect of the coronavirus (COVID-19) pandemic on health-related quality of life in children. Children and Youth Services Review 119, 105595.

2. Ahmed HO (2020) The impact of social distancing and self-isolation in the last corona COVID-19 outbreak on the body weight in Sulaimani governorate-Kurdistan/Iraq, a prospective case series study. Annals of Medicine and Surgery 59, 110–117. Elsevier.

3. Athanasiadis DI, Hernandez E, Hilgendorf W, et al. (2020) How are bariatric patients coping during the coronavirus disease 2019 (COVID-19) pandemic? Analysis of factors known to cause weight regain among postoperative bariatric patients. Surgery for Obesity and Related Diseases. Elsevier.

4. Blaszczyk-Bębenek E, Jagielski P, Bolesławska I, et al. (2020) Nutrition behaviors in Polish adults before and during covid-19 lockdown. Nutrients 12, 3084. Multidisciplinary Digital Publishing Institute.

5. Chagué F, Boulin M, Eicher J-C, et al. (2020) Impact of lockdown on patients with congestive heart failure during the coronavirus disease 2019 pandemic. ESC Heart Fail.

6. Chopra S, Ranjan P, Singh V, et al. (2020) Impact of COVID-19 on lifestyle-related behaviours-a cross-sectional audit of responses from nine hundred and ninety-five participants from India. Diabetes & Metabolic Syndrome: Clinical Research & Reviews 14, 2021–2030. Elsevier.

7. Cransac-Miet A, Zeller M, Chagué F, et al. (2021) Impact of COVID-19 lockdown on lifestyle adherence in stay-at-home patients with chronic coronary syndromes: Towards a time bomb. International Journal of Cardiology 323, 285–287. Elsevier.

8. Deschasaux-Tanguy M, Druesne-Pecollo N, Esseddik Y, et al. (2020) Diet and physical activity during the COVID-19 lockdown period (March-May 2020): results from the French NutriNet-Sante cohort study. medRxiv, 2020.06.04.20121855. Cold Spring Harbor Laboratory Press.

9. Di Santo SG, Franchini F, Filiputti B, et al. (2020) The effects of COVID-19 and quarantine measures on the lifestyles and mental health of people over 60 at increased risk of dementia. Frontiers in Psychiatry 11. Frontiers Media SA.

10. Di Renzo L, Gualtieri P, Pivari F, et al. (2020) Eating habits and lifestyle changes during COVID-19 lockdown: an Italian survey. Journal of Translational Medicine 18, 1–15. BioMed Central.

11. Dogas Z, Kalcina LL, Dodig IP, et al. (2020) The effect of COVID-19 lockdown on lifestyle and mood in Croatian general population: a cross-sectional study. Croatian medical journal 61, 309. Medicinska Naklada.

12. Dondi A, Candela E, Morigi F, et al. (2021) Parents’ Perception of Food Insecurity and of Its Effects on Their Children in Italy Six Months after the COVID-19 Pandemic Outbreak. Nutrients 13, 121. Multidisciplinary Digital Publishing Institute.

13. Dragun R, Veček NN, Marendić M, et al. (2021) Have Lifestyle Habits and Psychological Well-Being Changed among Adolescents and Medical Students Due to COVID-19 Lockdown in Croatia? Nutrients 13, 97. Multidisciplinary Digital Publishing Institute.

14. Drywień ME, Hamulka J, Zielinska-Pukos MA, et al. (2020) The COVID-19 Pandemic Lockdowns and Changes in Body Weight among Polish Women. A Cross-Sectional Online Survey PLifeCOVID-19 Study. Sustainability 12, 7768. Multidisciplinary Digital Publishing Institute.

15. de Matos DG, Aidar FJ, Almeida-Neto PF de, et al. (2020) The impact of measures recommended by the government to limit the spread of coronavirus (COVID-19) on physical activity levels, quality of life, and mental health of Brazilians. Sustainability 12, 9072. Multidisciplinary Digital Publishing Institute.

16. Gentile A, Torales J, O’Higgins M, et al. (2020) Phone-based outpatients’ follow-up in mental health centers during the COVID-19 quarantine. International Journal of Social Psychiatry, 0020764020979732. SAGE Publications Sage UK: London, England.

17. Giustino V, Parroco AM, Gennaro A, et al. (2020) Physical Activity Levels and Related Energy Expenditure during COVID-19 Quarantine among the Sicilian Active Population: A Cross-Sectional Online Survey Study. Sustainability 12, 4356. Multidisciplinary Digital Publishing Institute.

18. He M, Xian Y, Lv X, et al. (2020) Changes in Body Weight, Physical Activity, and Lifestyle During the Semi-lockdown Period After the Outbreak of COVID-19 in China: An Online Survey. Disaster Med Public Health Prep, 1–6.

19. Ismail LC, Osaili TM, Mohamad MN, et al. (2020) Assessment of Eating Habits and Lifestyle during Coronavirus Pandemic in the MENA region: A Cross-Sectional Study. British Journal of Nutrition, 1–30. Cambridge University Press.

20. Cheikh Ismail L, Osaili TM, Mohamad MN, et al. (2020) Eating habits and lifestyle during COVID-19 lockdown in the United Arab Emirates: a cross-sectional study. Nutrients 12, 3314. Multidisciplinary Digital Publishing Institute.

21. Jia P, Zhang L, Yu W, et al. (2020) Impact of COVID-19 lockdown on activity patterns and weight status among youths in China: the COVID-19 Impact on Lifestyle Change Survey (COINLICS). International Journal of Obesity, 1–5. Nature Publishing Group.

22. Jimenez A, de Hollanda A, Palou E, et al. (2021) Psychosocial, Lifestyle, and Body Weight Impact of COVID-19-Related Lockdown in a Sample of Participants with Current or Past History of Obesity in Spain. Obesity surgery, 1–10. Springer.

23. Kang HM, Jeong DC, Suh B-K, et al. (2021) The Impact of the Coronavirus Disease-2019 Pandemic on Childhood Obesity and Vitamin D Status. J Korean Med Sci 36. The Korean Academy of Medical Sciences.

24. Karatas S, Yesim T & Beysel S (2021) Impact of lockdown COVID-19 on metabolic control in type 2 diabetes mellitus and healthy people. Primary Care Diabetes. Elsevier.

25. Keel PK, Gomez MM, Harris L, et al. (2020) Gaining “The Quarantine 15:” Perceived versus observed weight changes in college students in the wake of COVID-19. International Journal of Eating Disorders 53, 1801–1808. Wiley Online Library.

26. Kriaucioniene V, Bagdonaviciene L, Rodríguez-Pérez C, et al. (2020) Associations between changes in health behaviours and body weight during the COVID-19 quarantine in Lithuania: the Lithuanian COVIDiet Study. Nutrients 12, 3119. Multidisciplinary Digital Publishing Institute.

27. Malkawi SH, Almhdawi K, Jaber AF, et al. (2020) COVID-19 Quarantine-Related Mental Health Symptoms and their Correlates among Mothers: A Cross Sectional Study. Maternal and Child Health Journal.

28. Marchitelli S, Mazza C, Lenzi A, et al. (2020) Weight Gain in a Sample of Patients Affected by Overweight/Obesity with and without a Psychiatric Diagnosis during the Covid-19 Lockdown. Nutrients 12, 3525. Multidisciplinary Digital Publishing Institute.

29. Martínez-de-Quel Ó, Suárez-Iglesias D, López-Flores M, et al. (2021) Physical activity, dietary habits and sleep quality before and during COVID-19 lockdown: A longitudinal study. Appetite 158, 105019.

30. Mason TB, Barrington-Trimis J & Leventhal AM (2021) Eating to Cope With the COVID-19 Pandemic and Body Weight Change in Young Adults. Journal of Adolescent Health 68, 277–283. Elsevier.

31. Mitchell ES, Yang Q, Behr H, et al. (2020) Self-reported food choices before and during COVID-19 lockdown. medRxiv, 2020.06.15.20131888. Cold Spring Harbor Laboratory Press.

32. Onmez A, Gamsızkan Z, Özdemir Ş, et al. (2020) The effect of COVID-19 lockdown on glycemic control in patients with type 2 diabetes mellitus in Turkey. Diabetes & Metabolic Syndrome: Clinical Research & Reviews 14, 1963–1966. Elsevier.

33. Ozden G & Parlar Kiliç S (2021) The Effect of Social Isolation during COVID-19 Pandemic on Nutrition and Exercise Behaviors of Nursing Students. Ecology of Food and Nutrition, 1–19. Taylor & Francis.

34. Pellegrini M, Ponzo V, Rosato R, et al. (2020) Changes in Weight and Nutritional Habits in Adults with Obesity during the ‘Lockdown’ Period Caused by the COVID-19 Virus Emergency. Nutrients 12.

35. Pietrobelli A, Pecoraro L, Ferruzzi A, et al. (2020) Effects of COVID-19 lockdown on lifestyle behaviors in children with obesity living in Verona, Italy: a longitudinal study. Obesity. Wiley Online Library.

36. Rogers N, Roberts C, Waterlow N, et al. (2020) Behavioural change towards reduced intensity physical activity is disproportionately prevalent among adults with serious health issues or self-perception of high risk during the UK COVID-19 lockdown. London School of Hygiene & Tropical Medicine.

37. Romero-Blanco C, Rodríguez-Almagro J, Onieva-Zafra MD, et al. (2020) Sleep Pattern Changes in Nursing Students during the COVID-19 Lockdown. Int J Environ Res Public Health 17.

38. Ruissen MM, Regeer H, Landstra CP, et al. (2021) Increased stress, weight gain and less exercise in relation to glycemic control in people with type 1 and type 2 diabetes during the COVID-19 pandemic. BMJ Open Diab Res Care 9, e002035.

39. Shah N, Karguppikar M, Bhor S, et al. (2020) Impact of lockdown for COVID-19 pandemic in Indian children and youth with type 1 diabetes from different socio-economic classes. Journal of Pediatric Endocrinology and Metabolism 1. De Gruyter.

40. Sidor A & Rzymski P (2020) Dietary Choices and Habits during COVID-19 Lockdown: Experience from Poland. Nutrients 12, 1657. Multidisciplinary Digital Publishing Institute.

41. Zachary Z, Brianna F, Brianna L, et al. (2020) Self-quarantine and Weight Gain Related Risk Factors During the COVID-19 Pandemic. Obesity Research & Clinical Practice. Elsevier.

## References

1. Deschasaux-Tanguy M, Druesne-Pecollo N, Esseddik Y, et al. (2020) Diet and physical activity during the COVID-19 lockdown period (March-May 2020): results from the French NutriNet-Sante cohort study. medRxiv, 2020.06.04.20121855. Cold Spring Harbor Laboratory Press.

2. Di Renzo L, Gualtieri P, Pivari F, et al. (2020) Eating habits and lifestyle changes during COVID-19 lockdown: an Italian survey. Journal of Translational Medicine 18, 1–15. BioMed Central.

3. Kriaucioniene V, Bagdonaviciene L, Rodríguez-Pérez C, et al. (2020) Associations between changes in health behaviours and body weight during the COVID-19 quarantine in Lithuania: the Lithuanian COVIDiet Study. Nutrients 12, 3119. Multidisciplinary Digital Publishing Institute.

4. Di Santo SG, Franchini F, Filiputti B, et al. (2020) The effects of COVID-19 and quarantine measures on the lifestyles and mental health of people over 60 at increased risk of dementia. Frontiers in Psychiatry 11. Frontiers Media SA.

5. Ozden G & Parlar Kiliç S (2021) The Effect of Social Isolation during COVID-19 Pandemic on Nutrition and Exercise Behaviors of Nursing Students. Ecology of Food and Nutrition, 1–19. Taylor & Francis.

6. Dogas Z, Kalcina LL, Dodig IP, et al. (2020) The effect of COVID-19 lockdown on lifestyle and mood in Croatian general population: a cross-sectional study. Croatian medical journal 61, 309. Medicinska Naklada.

7. Pellegrini M, Ponzo V, Rosato R, et al. (2020) Changes in Weight and Nutritional Habits in Adults with Obesity during the ‘Lockdown’ Period Caused by the COVID-19 Virus Emergency. Nutrients 12.

8. Pietrobelli A, Pecoraro L, Ferruzzi A, et al. (2020) Effects of COVID-19 lockdown on lifestyle behaviors in children with obesity living in Verona, Italy: a longitudinal study. Obesity. Wiley Online Library.

9. Sidor A & Rzymski P (2020) Dietary Choices and Habits during COVID-19 Lockdown: Experience from Poland. Nutrients 12, 1657. Multidisciplinary Digital Publishing Institute.

10. Jimenez A, de Hollanda A, Palou E, et al. (2021) Psychosocial, Lifestyle, and Body Weight Impact of COVID-19-Related Lockdown in a Sample of Participants with Current or Past History of Obesity in Spain. Obesity surgery, 1–10. Springer.

11. Marchitelli S, Mazza C, Lenzi A, et al. (2020) Weight Gain in a Sample of Patients Affected by Overweight/Obesity with and without a Psychiatric Diagnosis during the Covid-19 Lockdown. Nutrients 12, 3525. Multidisciplinary Digital Publishing Institute.

12. Drywień ME, Hamulka J, Zielinska-Pukos MA, et al. (2020) The COVID-19 Pandemic Lockdowns and Changes in Body Weight among Polish Women. A Cross-Sectional Online Survey PLifeCOVID-19 Study. Sustainability 12, 7768. Multidisciplinary Digital Publishing Institute.

13. Blaszczyk-Bębenek E, Jagielski P, Bolesławska I, et al. (2020) Nutrition behaviors in Polish adults before and during covid-19 lockdown. Nutrients 12, 3084. Multidisciplinary Digital Publishing Institute.

14. Mason TB, Barrington-Trimis J & Leventhal AM (2021) Eating to Cope With the COVID-19 Pandemic and Body Weight Change in Young Adults. Journal of Adolescent Health 68, 277–283. Elsevier.

15. Adıbelli D & Sümen A (2020) The effect of the coronavirus (COVID-19) pandemic on health-related quality of life in children. Children and Youth Services Review 119, 105595.

16. He M, Xian Y, Lv X, et al. (2020) Changes in Body Weight, Physical Activity, and Lifestyle During the Semi-lockdown Period After the Outbreak of COVID-19 in China: An Online Survey. Disaster Med Public Health Prep, 1–6.

17. Kang HM, Jeong DC, Suh B-K, et al. (2021) The Impact of the Coronavirus Disease-2019 Pandemic on Childhood Obesity and Vitamin D Status. J Korean Med Sci 36. The Korean Academy of Medical Sciences.

18. Rogers N, Roberts C, Waterlow N, et al. (2020) Behavioural change towards reduced intensity physical activity is disproportionately prevalent among adults with serious health issues or self-perception of high risk during the UK COVID-19 lockdown. London School of Hygiene & Tropical Medicine.

19. Malkawi SH, Almhdawi K, Jaber AF, et al. (2020) COVID-19 Quarantine-Related Mental Health Symptoms and their Correlates among Mothers: A Cross Sectional Study. Maternal and Child Health Journal.

21. Keel PK, Gomez MM, Harris L, et al. (2020) Gaining “The Quarantine 15:” Perceived versus observed weight changes in college students in the wake of COVID-19. International Journal of Eating Disorders 53, 1801–1808. Wiley Online Library.

23. Mitchell ES, Yang Q, Behr H, et al. (2020) Self-reported food choices before and during COVID-19 lockdown. medRxiv, 2020.06.15.20131888. Cold Spring Harbor Laboratory Press.

24. Athanasiadis DI, Hernandez E, Hilgendorf W, et al. (2020) How are bariatric patients coping during the coronavirus disease 2019 (COVID-19) pandemic? Analysis of factors known to cause weight regain among postoperative bariatric patients. Surgery for Obesity and Related Diseases. Elsevier.

25. Dragun R, Veček NN, Marendić M, et al. (2021) Have Lifestyle Habits and Psychological Well-Being Changed among Adolescents and Medical Students Due to COVID-19 Lockdown in Croatia? Nutrients 13, 97. Multidisciplinary Digital Publishing Institute.

27. Marchitelli S, Mazza C, Lenzi A, et al. (2020) Weight Gain in a Sample of Patients Affected by Overweight/Obesity with and without a Psychiatric Diagnosis during the Covid-19 Lockdown. Nutrients 12, 3525. Multidisciplinary Digital Publishing Institute.

28. Romero-Blanco C, Rodríguez-Almagro J, Onieva-Zafra MD, et al. (2020) Sleep Pattern Changes in Nursing Students during the COVID-19 Lockdown. Int J Environ Res Public Health 17.

29. Martínez-de-Quel Ó, Suárez-Iglesias D, López-Flores M, et al. (2021) Physical activity, dietary habits and sleep quality before and during COVID-19 lockdown: A longitudinal study. Appetite 158, 105019. Elsevier.

30. de Matos DG, Aidar FJ, Almeida-Neto PF de, et al. (2020) The impact of measures recommended by the government to limit the spread of coronavirus (COVID-19) on physical activity levels, quality of life, and mental health of Brazilians. Sustainability 12, 9072. Multidisciplinary Digital Publishing Institute.

31. Chopra S, Ranjan P, Singh V, et al. (2020) Impact of COVID-19 on lifestyle-related behaviours-a cross-sectional audit of responses from nine hundred and ninety-five participants from India. Diabetes & Metabolic Syndrome: Clinical Research & Reviews 14, 2021–2030. Elsevier.

32. Ahmed HO (2020) The impact of social distancing and self-isolation in the last corona COVID-19 outbreak on the body weight in Sulaimani governorate-Kurdistan/Iraq, a prospective case series study. Annals of Medicine and Surgery 59, 110–117. Elsevier.

33. Cransac-Miet A, Zeller M, Chagué F, et al. (2021) Impact of COVID-19 lockdown on lifestyle adherence in stay-at-home patients with chronic coronary syndromes: Towards a time bomb. International Journal of Cardiology 323, 285–287. Elsevier.

34. Shah N, Karguppikar M, Bhor S, et al. (2020) Impact of lockdown for COVID-19 pandemic in Indian children and youth with type 1 diabetes from different socio-economic classes. Journal of Pediatric Endocrinology and Metabolism 1. De Gruyter.

35. Giustino V, Parroco AM, Gennaro A, et al. (2020) Physical Activity Levels and Related Energy Expenditure during COVID-19 Quarantine among the Sicilian Active Population: A Cross-Sectional Online Survey Study. Sustainability 12, 4356. Multidisciplinary Digital Publishing Institute.

36. Jia P, Zhang L, Yu W, et al. (2020) Impact of COVID-19 lockdown on activity patterns and weight status among youths in China: the COVID-19 Impact on Lifestyle Change Survey (COINLICS). International Journal of Obesity, 1–5. Nature Publishing Group.

37. Ruissen MM, Regeer H, Landstra CP, et al. (2021) Increased stress, weight gain and less exercise in relation to glycemic control in people with type 1 and type 2 diabetes during the COVID-19 pandemic. BMJ Open Diab Res Care 9, e002035.

38. Ismail LC, Osaili TM, Mohamad MN, et al. (2020) Assessment of Eating Habits and Lifestyle during Coronavirus Pandemic in the MENA region: A Cross-Sectional Study. British Journal of Nutrition, 1–30. Cambridge University Press.

39. Chagué F, Boulin M, Eicher J-C, et al. (2020) Impact of lockdown on patients with congestive heart failure during the coronavirus disease 2019 pandemic. ESC Heart Fail.

40. Onmez A, Gamsızkan Z, Özdemir Ş, et al. (2020) The effect of COVID-19 lockdown on glycemic control in patients with type 2 diabetes mellitus in Turkey. Diabetes & Metabolic Syndrome: Clinical Research & Reviews 14, 1963–1966. Elsevier.

41. Karatas S, Yesim T & Beysel S (2021) Impact of lockdown COVID-19 on metabolic control in type 2 diabetes mellitus and healthy people. Primary Care Diabetes. Elsevier.

